# Unraveling Attributes of COVID-19 Vaccine Hesitancy and Uptake in the U.S.: A Large Nationwide Study

**DOI:** 10.1101/2021.04.05.21254918

**Authors:** Sean D. McCabe, E. Adrianne Hammershaimb, David Cheng, Andy Shi, Derek Shyr, Shuting Shen, Lyndsey D. Cole, Jessica R. Cataldi, William Allen, Ryan Probasco, Ben Silbermann, Feng Zhang, Regan Marsh, Mark A. Travassos, Xihong Lin

**Affiliations:** The How We Feel Project, USA; Department of Biostatistics, Harvard T.H. Chan School of Public Health, Boston, MA, USA; Center for Vaccine Development and Global Health, University of Maryland School of Medicine, Baltimore, MD, USA; Department of Pediatrics, University of Maryland School of Medicine, Baltimore, MD, USA; Department of Pediatrics, University of Colorado Anschutz Medical Campus; Adult and Child Consortium for Health Outcomes Research and Delivery Science, University of Colorado Anschutz Medical Campus and Children’s Hospital Colorado, Aurora, CO; Broad Institute of MIT and Harvard, Cambridge, MA, USA; Society of Fellows, Harvard University, Cambridge, MA, USA; Department of Biological Engineering, Massachusetts Institute of Technology, Cambridge, MA, USA; McGovern Institute for Brain Research, Massachusetts Institute of Technology, Cambridge, MA, USA; Department of Brain and Cognitive Sciences, Massachusetts Institute of Technology, Cambridge, MA, USA; Howard Hughes Medical Institute, Chevy Chase, MD, USA; Department of Emergency Medicine, Brigham and Women’s Hospital, Boston, MA, USA; Department of Emergency Medicine, Harvard Medical School, Boston, MA, USA; Partners in Health, Boston, MA, USA; Department of Statistics, Harvard University, Cambridge, MA, USA

## Abstract

SARS-CoV-2 vaccines are powerful tools to combat the COVID-19 pandemic, but vaccine hesitancy threatens these vaccines’ effectiveness. To address COVID-19 vaccine hesitancy and ensure equitable distribution, understanding the extent of and factors associated with vaccine acceptance and uptake is critical. We report the results of a large nationwide study conducted December 2020-May 2021 of 34,470 users from COVID-19-focused smartphone-based app How We Feel on their willingness to receive a COVID-19 vaccine. Nineteen percent of respondents expressed vaccine hesitancy, the majority being undecided. Of those who were undecided or unlikely to get a COVID-19 vaccine, 86% reported they ultimately did receive a COVID-19 vaccine. We identified sociodemographic and behavioral factors that were associated with COVID-19 vaccine hesitancy and uptake, and we found several vulnerable groups at increased risk of COVID-19 burden, morbidity, and mortality were more likely to be vaccine hesitant and had lower rates of vaccination. Our findings highlight specific populations in which targeted efforts to develop education and outreach programs are needed to overcome vaccine hesitancy and improve equitable access, diversity, and inclusion in the national response to COVID-19.

---

The emergence in late 2019 of severe acute respiratory syndrome coronavirus 2 (SARS-CoV-2) as a novel human pathogen and causative agent of the global coronavirus disease 2019 (COVID-19) pandemic^1^ fueled an unprecedented effort to rapidly develop a vaccine^2^. While the successful development of several effective SARS-CoV-2 vaccines has given many hope, the defining challenge now to effectively combat COVID-19 is ensuring equitable vaccine distribution and high vaccine uptake.

At least 70% of the U.S. population needs immunity to SARS-CoV-2 to end the COVID-19 pandemic^3^. Public opinion polling in early 2020 suggested that as many as 72% of U.S. adults were willing to receive a COVID-19 vaccine once licensed and available. Four months later, the number of U.S. adults willing to receive a SARS-CoV-2 vaccine had sharply declined to as low as 51%^4^.

In December 2020, two vaccines against COVID-19 received Emergency Use Authorization (EUA) from the U.S. Food and Drug Administration^5,6^. The results of phase 3 clinical trials and the subsequent rollout of the Pfizer-BioNTech and Moderna vaccines received significant attention in the media. Opinion polls conducted in December 2020 suggested a subsequent increase in public willingness to receive a COVID-19 vaccine, likely due to the widespread availability of data showing the vaccines to be both safe and effective^7^. Despite Johnson & Johnson’s Janssen COVID-19 vaccine also receiving EUA, national uptake of vaccines has declined since mid-April as those reluctant to be vaccinated occupy a greater percentage of the unvaccinated population^8-10^.

In this paper, we associate the degree of COVID-19 vaccine hesitancy in the U.S. with characteristics that might influence vaccine acceptance and with eventual COVID-19 vaccine uptake. This will help public health and community leaders develop effective education and outreach programs to overcome vaccine hesitancy and ensure equitable vaccine distribution and improve vaccine uptake.

How We Feel (HWF; http://www.howwefeel.org) is a web and mobile-phone application developed to facilitate the large-scale collection of data about COVID-19 symptoms, SARS-CoV-2 test results, and transmission-mitigating behaviors and sentiments^11^ [8]. Users are assigned a randomly generated number that tracks logins from the same device and are otherwise unidentifiable. Beginning in December 2020, we fielded a question about users’ COVID-19 vaccine intentions: “If a safe, effective coronavirus vaccine were available, how likely would you be to get yourself vaccinated?” Responses were recorded on a 5-point bipolar Likert scale, and vaccine hesitancy was defined as a “Very Unlikely,” “Unlikely,” or “Undecided” response. These responses were then related to the user’s subsequent COVID-19 vaccine uptake or refusal. Users were asked “Have you received a COVID-19 vaccine?” and could respond with “Yes,” “No, I haven’t been offered one,” or “No, I have been offered one but declined.”

## Results

A total of 36,711 users responded to the vaccine intention question. The largest number of respondents came from Connecticut and California with 8,697 and 4,668, respectively (Supplementary Figure 1a). HWF’s user base is approximately 79% female (Supplementary Figure 1b) and 83% white (Supplementary Figure 1c). Users are 18 years of age or older and are equally distributed by age groups (Supplementary Figure 1d). More than 68% of respondents were non-essential workers, and users cover a diverse range of income groups. All descriptive statistics of the study participants are available in Supplementary Table 1.

In total, 30,618 (83%) were accepting (“Likely” or “Very Likely”) of the vaccine (Figure 1a). After applying a census-based post-stratification weight (see Methods), Vermont (92%) and Washington D.C. (88%) had the highest rates of vaccine acceptance while South Dakota (27%) and Louisiana (23%) had the highest rates of undecided users (Figure 1b). Weighted bar plots of vaccine intent across demographic characteristics revealed that “Undecided” users represented the largest proportion of hesitant users across all demographic groups (Figure 1c, Supplementary Table 2). State level hesitancy rates were negatively associated with the average number of users that practiced transmission mitigating behaviors and were positively associated with cumulative COVID-19 case and death rates by January 10, 2021 (Figure 2a). Unweighted plots are available in Supplementary Figure 2.

**Figure 1.**
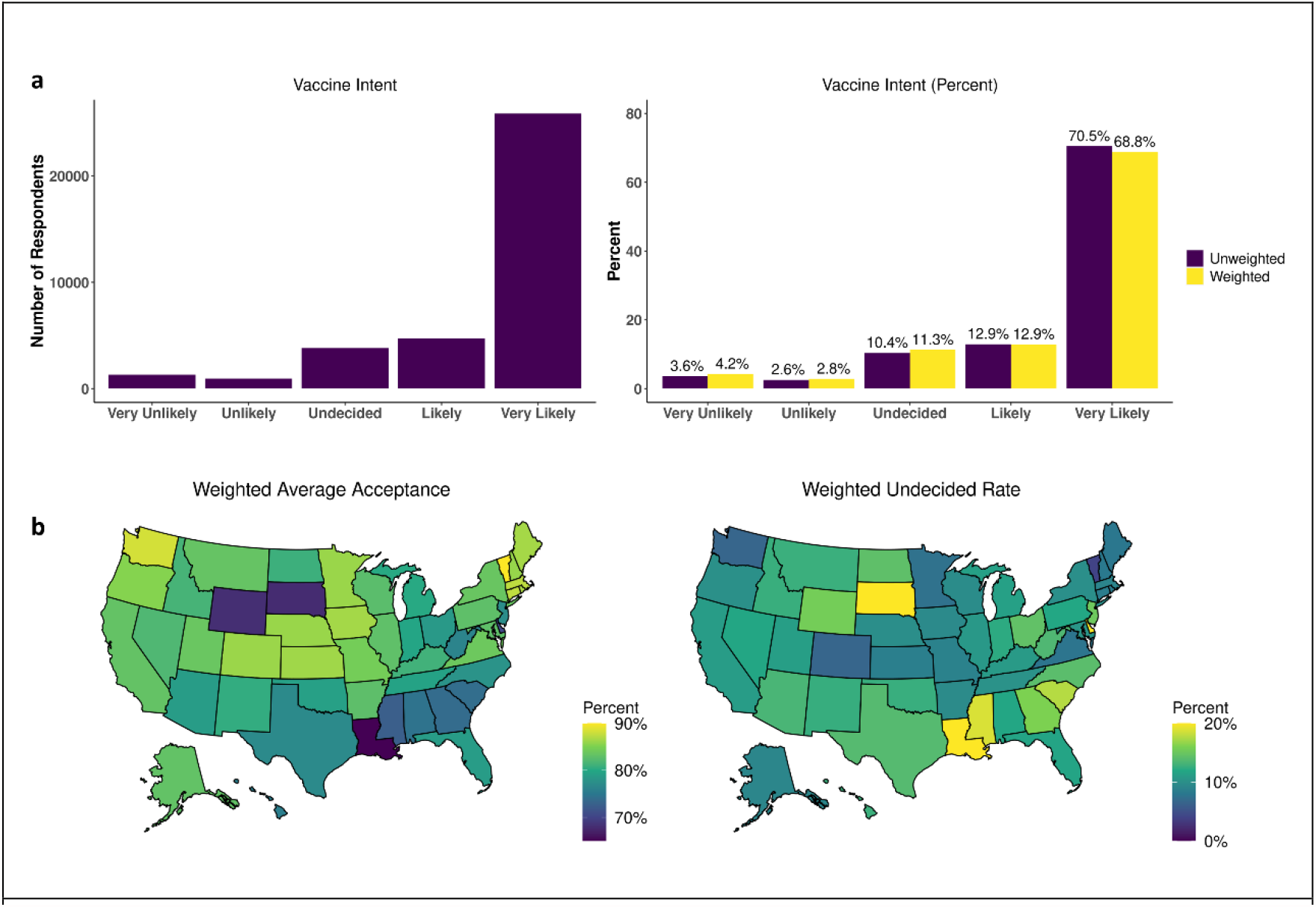
COVID-19 Hesitancy rates: **(a)** (Left) Number of responses and (Right) unweighted and weighted percentages. **(b)** Weighted average acceptance and undecided rates by state

**Figure 2.**
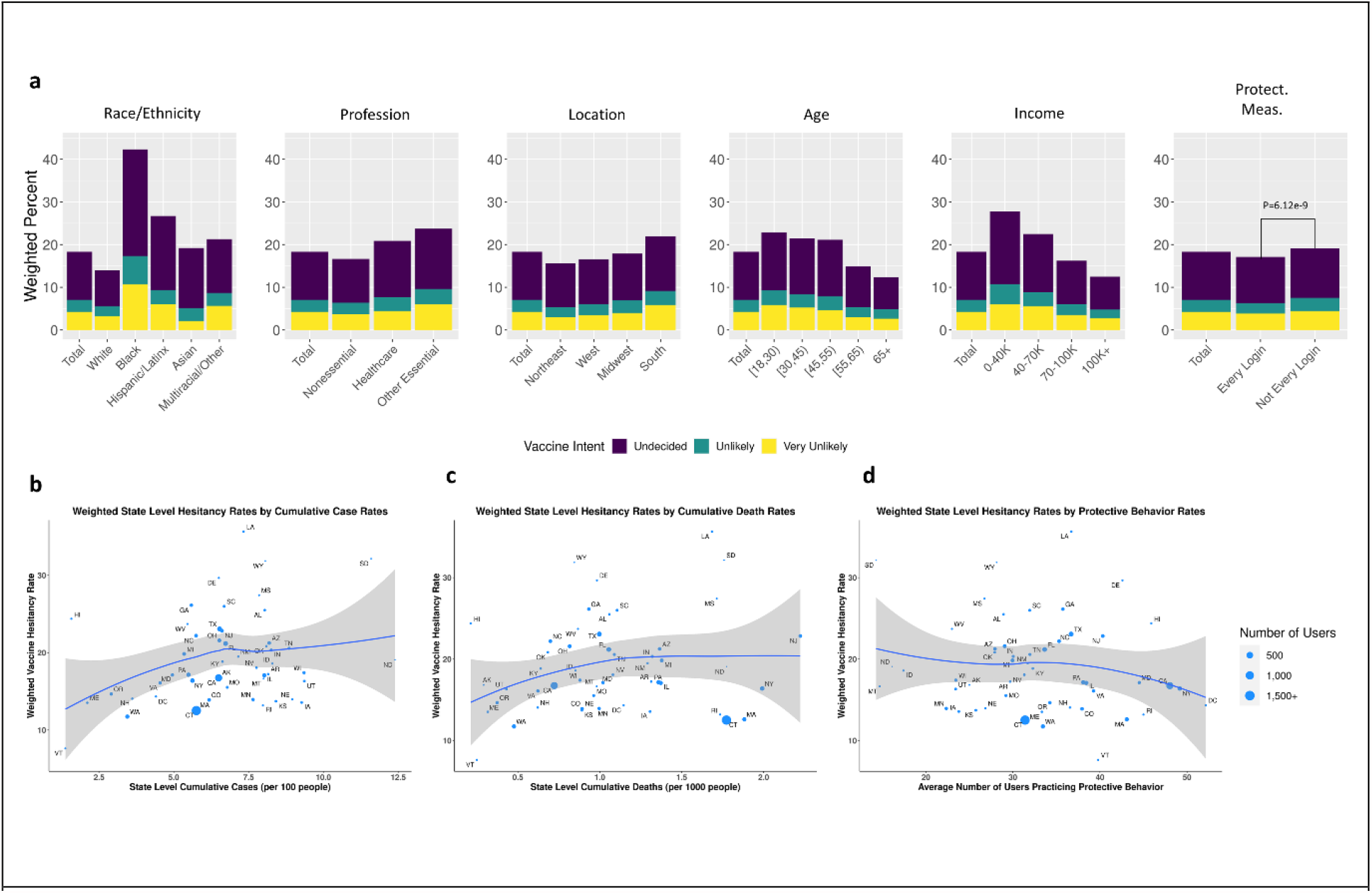
Demographic Hesitancy Rates: **(a)** Weighted percentages of hesitant responses by race/ethnicity, profession, location, age, income, and use of protective measures. State level weighted hesitancy rates by **(b)** cumulative case rates (/100 individuals), **(c)** cumulative death rates (/1000 individuals), **(d)** and average number of users practicing protective behavior.

To assess demographic associations with vaccine hesitancy, we fit a univariate logistic regression with socio-demographic, occupation, preexisting medical conditions, geographical and COVID-19 related predictors (Supplementary Tab 3) and a multivariable logistic regression model to adjust for potential confounding between the predictors (Figure 3, Supplementary Table 4). We implemented post-stratification weights using census estimates of sex, age, race, and census location (see Methods). People of color reported higher rates of vaccine hesitancy compared to white non-Hispanic users (African American OR, 3.94; CI, 3.47, 4.48; p=1.26e-96). Vaccine hesitancy was more likely among females than males (OR,1.67; CI, 1.51, 1.83; p=4.09e-25); younger users than those over 65 years old (18-30 OR, 2.17; CI, 1.86, 2.53; p=1.03e-22); those with three or more preexisting conditions than those with zero (OR, 1.19; CI, 1.06, 1.34; p=0.0036); and parents than non-parents (OR, 1.26; CI, 1.15, 1.38; p=9.61e-7).

**Figure 3.**
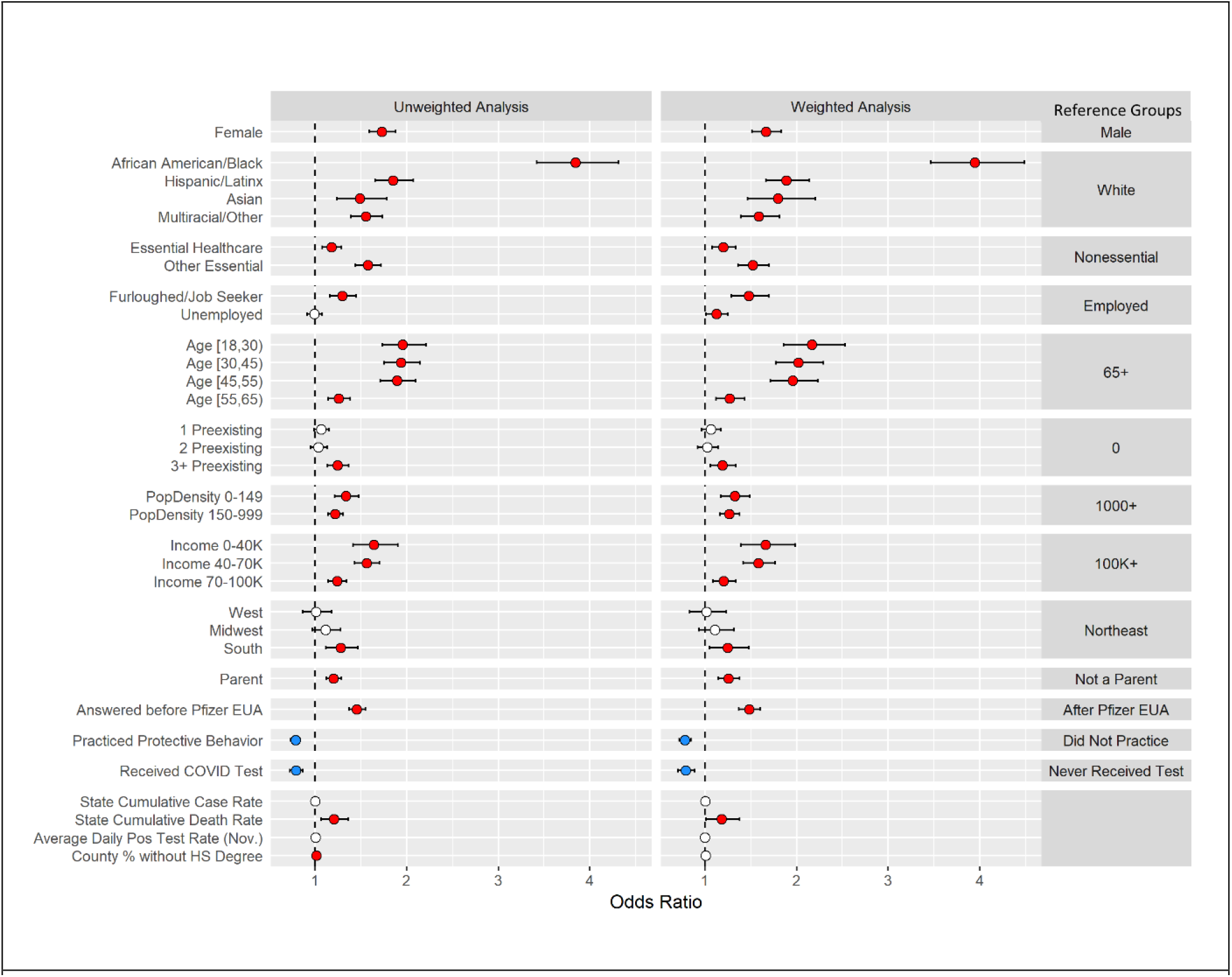
Logistic regression-based association analysis results of vaccine hesitancy: Forest plots for (Left) unweighted and (Right) weighted multivariable logistic regression analyses for vaccine hesitancy with 95% confidence intervals. Non-significant variables at the 0.05 level (white), significant positive associations (red), and significant negative associations (blue).

Individuals that were furloughed or job-seeking were also more vaccine hesitant compared to those working full- or part-time (OR, 1.48; CI, 1.29, 1.70; p=4.04e-8). Respondents from the South (OR, 1.25; CI, 1.05, 1.48; p=0.0105), from less densely populated areas, or with lower incomes were all more likely to be vaccine hesitant. Users that responded before the Pfizer Emergency Use Authorization (EUA) on December 11, 2020 were more vaccine hesitant than users who responded after the Pfizer EUA (OR, 1.48; CI, 1.37, 1.60; p=9.96e-23), users who practiced behavior protective against COVID-19 such as mask-wearing or social distancing were less vaccine hesitant (OR, 0.78; CI, 0.72, 0.85; p=6.12e-9), and users that received a COVID test were less vaccine hesitant (OR, 0.79; CI, 0.71, 0.89, p=5.88e-5).

Nominal logistic regression (see Methods) evaluated whether vaccine hesitancy was driven by “Undecided” vs. “Unlikely/Very Unlikely” responses (Supplementary Table 5), and was also conducted with a weighted analysis (Supplementary Table 6). Hesitancy in healthcare workers, those aged 55-65, Asian users, and those in locations with a median income between $70,000 and $100,000 was driven by the “Undecided” group, whereas hesitancy in the unemployed, those with 3+ preexisting conditions, and southern users was driven by the “Unlikely” group. Sensitivity analyses were performed for the weighted multivariable and nominal regression analyses with a less restrictive threshold for the trimming weights (Supplementary Table 7/8) and found similar results. We conducted a sensitivity analyses to assess differences in hesitancy in individuals that tested positive for COVID-19 and found no difference in intention based on testing results (see Methods, Supplementary Table 9/10).

Of the 36,711 users who responded to the vaccine intent question, 23,429 also responded to the vaccine uptake question with 98% (18,230/18,680) of users who were offered a COVID-19 vaccine accepting vaccination (Figure 4a). Demographic distributions remained similar to those of respondents of the vaccine intent question with a slight increase in the proportion of users ages 55+ (Supplementary Figure 3). Vaccination rates by state are shown in Figure 4b for all users that responded to the vaccine uptake question and subset to respondents who were offered a vaccine. Users with lower levels of vaccine intent had lower rates of vaccination (Figure 4c), and Black and Hispanic/Latinx users reported lower rates of vaccination than White, Non-Hispanic users (Figure 4d). Plots of weighted and unweighted vaccination rates across all demographic features are available in Supplementary Figures 4-5.

**Figure 4.**
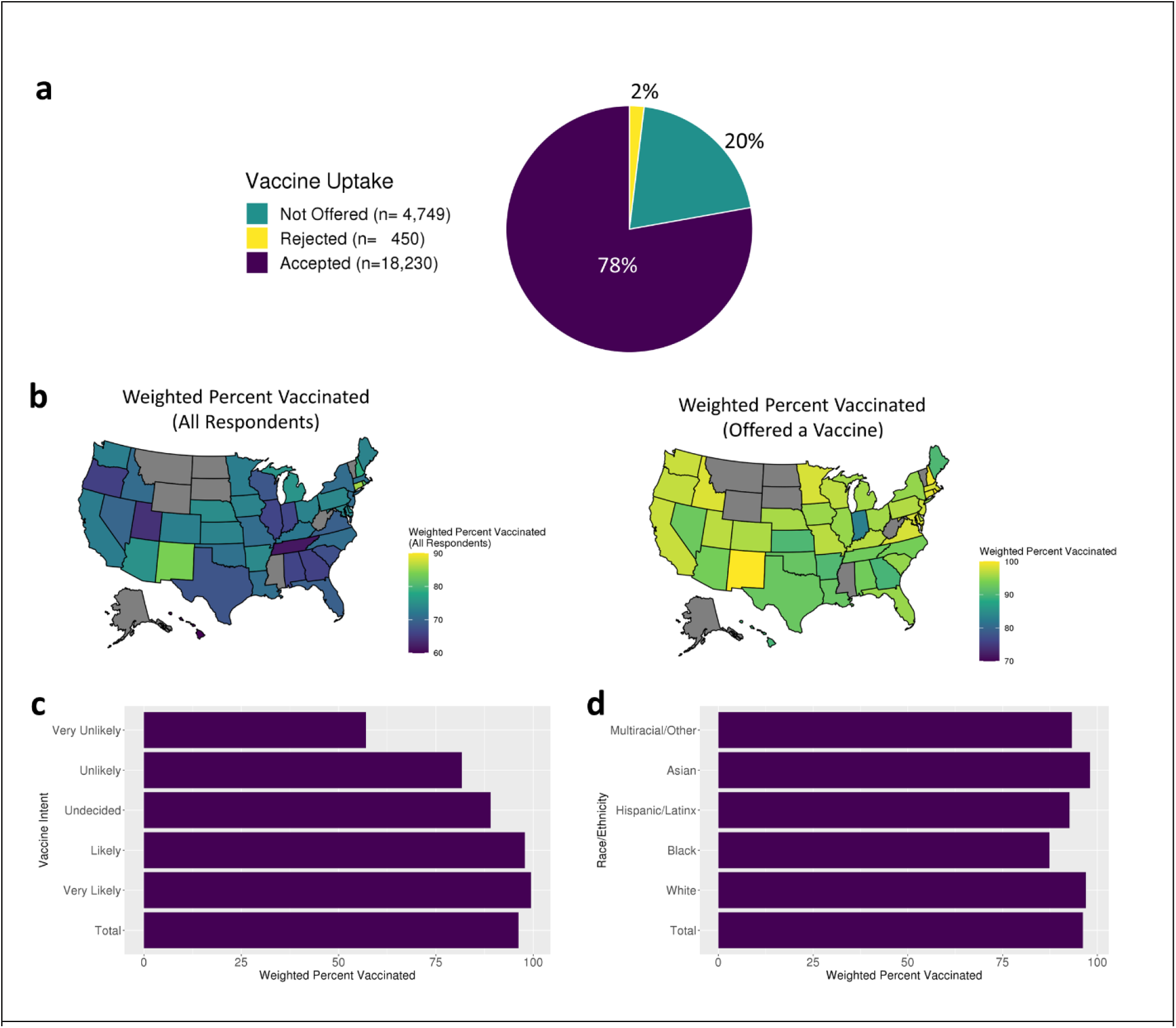
Vaccine Uptake Rates: **(a)** Vaccine uptake responses for all users. **(b)** Weighted vaccination rates by state of (Left) all users that responded to the vaccine uptake question and (right) users that were offered a vaccine. **(c)** Weighted vaccination rates of users that were offered a vaccine by vaccine intent and **(d)** race/ethnicity.

To formally identify demographic features associated with differences in vaccination rates, we conducted an unweighted and weighted multiple logistic regression analysis (see Methods, Figure 5, Supplementary Table 11/12). All age groups reported lower rates of vaccinations compared to users over 65 (18-30 OR: 0.10; CI, 0.06, 0.18; p=1.43e-16); Black users reported lower rates of vaccinations (OR, 0.58; CI, 0.38, 0.91; p=0.0165) compared to White non-Hispanic users; essential workers outside of healthcare reported lower rates of vaccinations (OR, 0.64; CI, 0.44, 0.92; p=0.0162) compared to non-essential workers; parents reported lower rates of vaccination (OR, 0.63; CI, 0.45, 0.89; p=0.0086) compared to users who are not parents; users in areas with a median household income (MHI) of $40-70K (OR, 0.56; CI, 0.37, 0.85; p=0.0066) and $70-100K (OR, 0.63; CI, 0.42, 0.96; p=0.0316) reported lower rates of vaccinations compared to those in areas with a MHI $100K+; users logging in from areas with 0-149 people/sq. mi reported lower rates of vaccinations (OR, 0.53; CI, 0.34, 0.82; p=0.0049) compared to users in high population density areas; and users that responded “Unlikely/Very Unlikely” (OR, 0.02; CI, 0.01, 0.03; p=2.07e-114) and “Undecided” (OR, 0.08; CI, 0.06, 0.12; p=1.06e-39) to the vaccine intent question reported lower rates of vaccinations compared to non-hesitant users.

**Figure 5.**
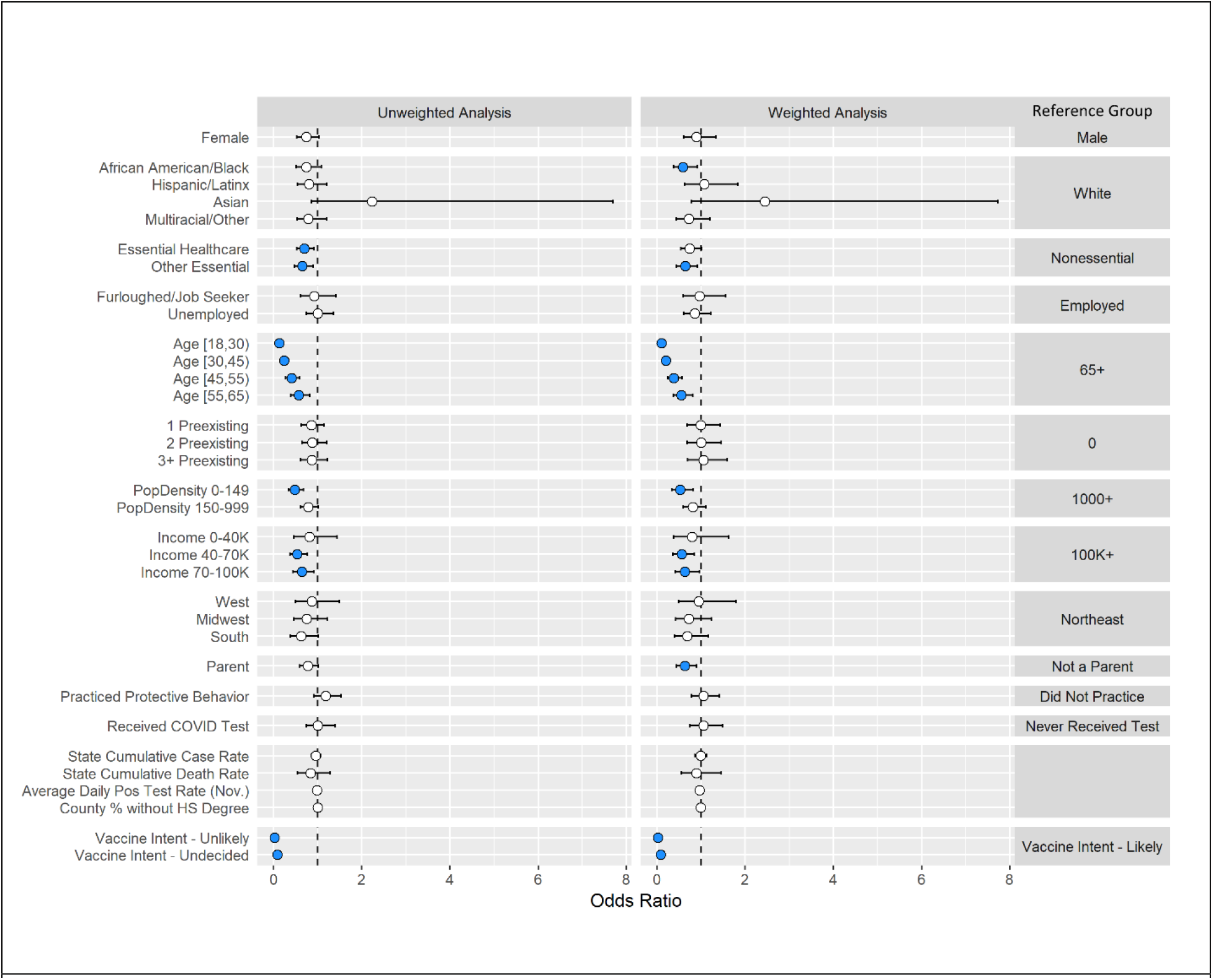
Logistic regression-based association analysis results of vaccine uptake: Forest plots for (Left) unweighted and (Right) weighted multivariable logistic regression analyses for vaccination uptake with 95% confidence intervals. Non-significant variables at the 0.05 level (white), significant positive associations (red), and significant negative associations (blue).

While vaccination rates were lower in the hesitant group compared to the non-hesitant group, 86% (2,157/2,520) of hesitant users were vaccinated. In a formal multiple regression analysis looking at demographic associations with vaccine uptake among hesitant users, similar associations were found (see Methods, Supplementary Table 13). Younger age groups, healthcare workers, people from lower income households, and residents of areas with lower population density had lower vaccination rates. Users who responded to the vaccine intent question as “Undecided” reported higher rates of vaccination compared to those that responded “Unlikely/Very Unlikely” (OR, 4.57; CI, 3.47, 6.03; p=2.26e-26).

## Discussion

Nationwide, we found lower rates of vaccine hesitancy than previously estimated, potentially related to confidence in the COVID-19 vaccines now available. Increased hesitancy was associated with minority race/ethnicity, living in less densely populated regions, and being a healthcare worker. A large proportion of these populations were undecided about COVID-19 vaccination, suggesting that targeted outreach may improve vaccine uptake. In fact, a significant portion of those skeptical or undecided about vaccination were ultimately vaccinated, supporting the idea that perspectives on COVID-19 vaccination are not immutable and may respond to such outreach.

Black respondents had the highest rates of COVID-19 vaccine hesitancy and the lowest rates of vaccine uptake relative to other racial and ethnic groups, consistent with other surveys^4, 9,10, 12-15^. The history of racist practices within the U.S. healthcare system and research community, such as during the Tuskegee Syphilis Study^16^, and disparities in social determinants of health including poor access to healthcare and limited time off work likely contribute to our findings. Dispelling concerns within the Black community requires extensive, sustained, structured outreach and will be critical to efforts to contain and eliminate COVID-19. The National Institutes of Health’s Community Engagement Alliance (CEAL) provides a model for such outreach, targeting populations that have been hardest hit by the COVID-19 pandemic^17^.

Education and outreach efforts must target several additional populations. This includes rural residents and young adults. Because large proportions of these populations were undecided about COVID-19 vaccination, outreach to these groups must also provide reliable vaccine information tailored to the needs of each community, and different outreach strategies may be needed to address the concerns of those who were undecided and those who were unlikely.

Vaccine hesitancy in healthcare workers warrants particular attention. We found that hesitant healthcare workers were less likely to change their mind than other hesitant workers (Supplementary Table 13), and previous work found that U.S. nurses had the highest degree of COVID-19 vaccine hesitancy among healthcare workers^18^. As the profession that enjoys the highest degree of public trust, nurses have an important role to play in promoting vaccine confidence^19^. Furthermore, inadequate vaccine uptake among healthcare workers raises the possibility of sustained COVID-19 transmission in an essential worker population critical to caring for vulnerable members of society, including immunocompromised individuals and children, the majority of whom were not yet eligible for a COVID-19 vaccine as of June 15, 2021^20-23^.

Addressing regional foci of vaccine hesitancy will be critical in federal resource allocation to combat vaccine hesitancy. We identified the greatest level of COVID-19 vaccine hesitancy in the South followed by the Midwest. While a CDC-sponsored survey conducted in December 2020 found that hesitancy was most prevalent in the Northeast, followed by the South^15^, other data from the CDC detailing U.S. state and county-level vaccination rates and allocated dose usage have consistently shown that Southern states have lower vaccination rates and lower allocated dose usages compared to other areas of the country^9^.

Initial reluctance or indecision regarding COVID-19 vaccination was not fixed and did not necessarily reflect a respondent’s eventual vaccination decision. This suggests the need for a multi-pronged approach that includes interventions directed at behavior change and that importantly is not discouraged by high rates of vaccine hesitancy. Even if receptivity towards vaccination is low, there may still be significant potential for increasing vaccine uptake, indicating the need for continued implementation of strategies known to be effective, such as health care provider outreach and reminders^24,25^.

A study limitation is that our sample may not be generalizable to the broader American public. How We Feel users are self-selecting and more likely to have a baseline level of concern about COVID-19. The user base is inherently skewed by a large proportion of users residing in Connecticut and California and by regional age discrepancies. Census-adjusted, weighted analysis help correct the sampling bias but may not completely remove the potential for bias, and interpretation of our findings should note this. Additionally, we were unable to objectively verify self-reported vaccination; however, studies of influenza vaccination have shown high degrees of concordance between self-reported vaccination and respondents’ actual vaccination status^26,27^.

Further work is needed to better understand how vaccine hesitancy relates to novel vaccine uptake in the U.S. and to understand how knowledge, attitudes, and behaviors surrounding COVID-19 vaccines change over time. As COVID-19 vaccines have become widely available to adults and adolescents in the U.S. and COVID-19 restrictions are lifting, our findings affirm the ongoing need to address vaccine hesitancy and issues related to access.

## Supporting information

STROBE Checklist

## Data Availability

This work used data from the How We Feel project (http://www.howwefeel.org/). The data are not publicly available, but researchers can apply to use the resource. Researchers with an appropriate IRB approval and data security approval to perform research involving human subjects using the HowWeFeel data can apply to obtain access to data used in the analysis.

## Online Methods

### Ethics statement

The HWF application was approved as exempt by the Ethical & Independent Review Services LLP IRB (Study ID 20049–01). The analysis of HWF data was also approved as exempt by Harvard University Longwood Medical Area Institutional Review Board (IRB) (Protocol no. IRB20-0514) and the Broad Institute of MIT and Harvard IRB (Protocol no. EX-1653). Informed consent was obtained from all users and the data were collected in de-identified form.

### Open-source software

We used the following open-source software in the analysis.

- R: http://www.r-project.org
- Tidyverse: http://www.tidyverse.org
- Data.table: https://CRAN.R-project.org/package=data.table
- nnet https://CRAN.R-project.org/package=nnet
- censusapi https://CRAN.R-project.org/package=censusapi
- survey https://CRAN.R-project.org/package=survey
- ggplot2 https://CRAN.R-project.org/package=ggplot2
- cowplot https://CRAN.R-project.org/package=cowplot

### Data Collection

Data were collected from the How We Feel web and mobile application on vaccine hesitancy between December 4^th^, 2020 and May 6^th^ 2021. Users were asked “If a safe, effective coronavirus vaccine were available, how likely would you be to get yourself vaccinated?” Responses were given on a bipolar 5-point Likert scale from “Very Unlikely” to “Very Likely”, with “Undecided” being the middle value. Users were asked the vaccine hesitancy question at regular intervals and the first response was used for the analysis. On February 12^th^, 2021, a vaccine uptake question was added. Users were asked “Have you received a COVID-19 vaccine?” and could respond with “Yes”, “No, I haven’t been offered one”, or “No, I have been offered one but declined”. For all uptake models the most recent response was used.

Users also self-reported race/ethnicity, sex, age, occupation, and preexisting conditions. Users who identified as “other” in the gender response were dropped due to small sample size. Neighborhood specific median household income was obtained from the user’s zip code by using the American Community Survey 5-year average results from 2018. Population density was calculated at the county level for each user based on data from the Yu Group at University of California at Berkeley^28^. State level case and death rates were obtained from USAFACTS^29^.

Race/ethnicity was defined using distinct groups corresponding to “White,” “Black/African-American,” “Hispanic/Latino,” and “Asian” if the user only selected that respective racial group. Users which answered more than one race or ethnicity or selected an option other than the ones listed above were placed in a “multiracial/other” category.

During each login, users reported whether they left their home and for what reason. If they left home, they were then asked what types of protective measurements they used while away (mask, social distancing, cloth mask, and/or avoiding public transportation). We defined “protective behavior” to be if a user either stayed home or wore a mask when outside the home. If the user said that they did not wear a mask outside the home but engaged only in outdoor exercise and maintained physical distance from others, then they were also considered to be practicing protective behavior. We then created a variable that was coded as “1” if they practiced protective behavior during all logins and a “0” if they failed to be protective during at least one login.

### Modeling

Users were considered to be vaccine hesitant if they responded “Very Unlikely,” “Unlikely,” or “Undecided” to the vaccine question. Using vaccine hesitancy as the outcome, a logistic regression was fit using several demographic variables as predictors to identify characteristics of users that were more or less vaccine hesitant. Both a univariate (Supplementary Table 3) and a multivariable model (Figure 3, Supplementary Tab 4) were performed to adjust for potential confounding. Only responses from users residing within the United States were used in the modelling. Corresponding odds ratios and 95% confidence intervals are provided, and statistical significance was assessed at the 0.05 level. Analyses were conducted using R (v 3.5.1).

Using the same covariates as in the logistic regression, a nominal logistic regression was fit to assess if results from the logistic regression were driven by individuals being more likely to be in the “Undecided” or “Unlikely” groups. The 5-point Likert scale was reduced to a 3-level bipolar variable for modelling purposes by combining “Very Unlikely” with “Unlikely” and “Very Likely” with “Likely” (Supplementary Table 5).

### Weighted Analysis

To adjust our analyses to a user base that matches the major U.S. census demographics, we implemented a weighted analysis using post-stratification weights. Using the census population estimates of sex, race, age, and census location, a population-based joint distribution was obtained. A user base distribution was also calculated using the same breakdown, and the two proportions were then matched per user. The post-stratification weight was then calculated by dividing the census proportion by the sample proportion plus 1e-4 to avoid issues with smaller user base probabilities. To avoid over or underweighting individuals, the post-stratification weights were trimmed to be between 0.3 and 3 using the *trimWeights* function in the *survey* R package. The weighted analysis was then conducted using the *svyglm* function (Supplementary Table 4). For the nominal regression analysis, two separate weighted logistic regressions were conducted. One compared the “Undecided” group vs. the “Likely” group, while the other compared the “Unlikely” group vs. the “Likely” group (Supplementary Table 6). To assess the choice of the weight trimming bounds, sensitivity analyses were conducted for both of the above weighted analyses (Supplementary Table 7/8) using a threshold of 0.1 and 5. Supplementary Figure 6 provides the distribution of the post-stratification weights.

### IPW Analysis

To formally assess if there was a difference in vaccine hesitancy between those that received a prior positive COVID test and those that received a negative test, we adjust for the demographic biases associated with receiving a COVID test. We first fit a weighted logistic regression to model the probability of receiving a test using all individuals and all demographic features that have been reported in previous analyses while applying the same weighted procedure as above. The coefficients, 95% confidence intervals, and p-values for this analysis are available in Supplementary Table 9. The fitted probabilities were then used as inverse probability weights (IPWs) in a weighted logistic regression model for vaccine hesitancy only including individuals which had received a COVID test. The same predictors for previous weighted models were used and a new variable designating if a user tested positive or negative was included. To avoid extreme high or low weights, the fitted probabilities were trimmed to be between 0.1 and 0.9 or 0.05 and 0.95. The results of both of these models are available in Supplementary Table 10.

### Vaccine Uptake

An unweighted multivariable logistic regression model was fit to identify which demographic features were associated with accepting or rejecting a COVID-19 vaccine. Along with the covariates included in the vaccine intent model, the three-level vaccine intent variable (“Very Likely/Likely”, “Undecided”, “Very Unlikely/Unlikely”) was also included in the analysis. Results are available in Supplementary Table 12 (left). To account for the biased sampling, non-response bias, and demographic differences in being offered a vaccine, a weighted multivariable model was fit. First, a weighted multivariable logistic regression model was fit for the probability of an individual responding to the vaccine uptake question with the inclusion of post-stratification weights as was done in the weighted vaccine intent model (Supplementary Table 11 left). The fitted probabilities from this model were then used as inverse probability weights to model the probability of a user being offered a vaccine (Supplementary Table 11 right). A user was defined as being offered a vaccine if the user responded to the question “Have you received a COVID-19 vaccine?” with “Yes,” or “No, I have been offered one but declined,” compared to users responding “No, I have not been offered a vaccine.” The fitted probabilities from this model were multiplied by the fitted probabilities from the response model and used as inverse probability weights in a final model which models the probability of accepting or rejecting the vaccine. The coefficients, 95% confidence intervals, and p-values for the final weighted model are available in Supplementary Table 12. To more formally characterize the attributes associated with vaccine uptake within users that responded as vaccine hesitant, we fit a weighted multivariable logistic regression model subset to only the users who initially responded they were “Very Unlikely” or “Unlikely” to receive a COVID-19 vaccine. Models were fit identically to the above weighted models for all users and results of the final model are available in Supplementary Table 13.

## Acknowledgements

S.D.M. is supported by the United States National Institutes of Health [grant T32ES007069] and a grant from the Partners in Health during preparation and writing of this manuscript. E.A.H. is supported by the United States National Institutes of Health [grant T32A1007524] during preparation and writing of this manuscript. X.L. is supported by a grant from the Partners in Health. D.S. is supported by United States National Institutes of Health [grant T32GM135117]. F.Z. is supported by the Howard Hughes Medical Institute, the McGovern Foundation, and J. and P. Poitras and the Poitras Center. The How We Feel Project is a non-profit corporation. The How We Feel Project thanks many operational volunteers and the HWF participants who took our survey and allowed us to share our analysis. Funding and in-kind donations for the How We Feel Project came from B. and D. Silbermann, F. Zhang and Y. Shi, L. Harp McGovern, D. Cheng, A. Azhir, K.H. Yoon and the Bill & Melinda Gates Foundation.

## Code Availability

The analysis code developed for this paper can be found online at https://github.com/mccabes292/HWF_VaccineHes_Paper.

## Contributions

E.A.H. and S.D.M. initiated the project. S.D.M. led data analysis and figure production. S.D.M., D. S. and S. S. cleaned the data. E.A.H., and A.S. contributed to analysis. R. M. provided feedback on analysis. S.D.M. and E.A.H. wrote the manuscript with M.T. and X.L. D. C., W. A., R.P., B.S., F. Z., X. L. designed and implemented the How We Feel application. E.A.H., L.D.C., J.R.C., and R.M. developed the vaccine hesitancy and uptake instrument. M.T. and X.L. supervised all aspects of the work.

