## Supplementary Figures for "Unraveling Attributes of COVID-19 Vaccine Hesitancy and Uptake in the U.S.: A Large Nationwide Study"

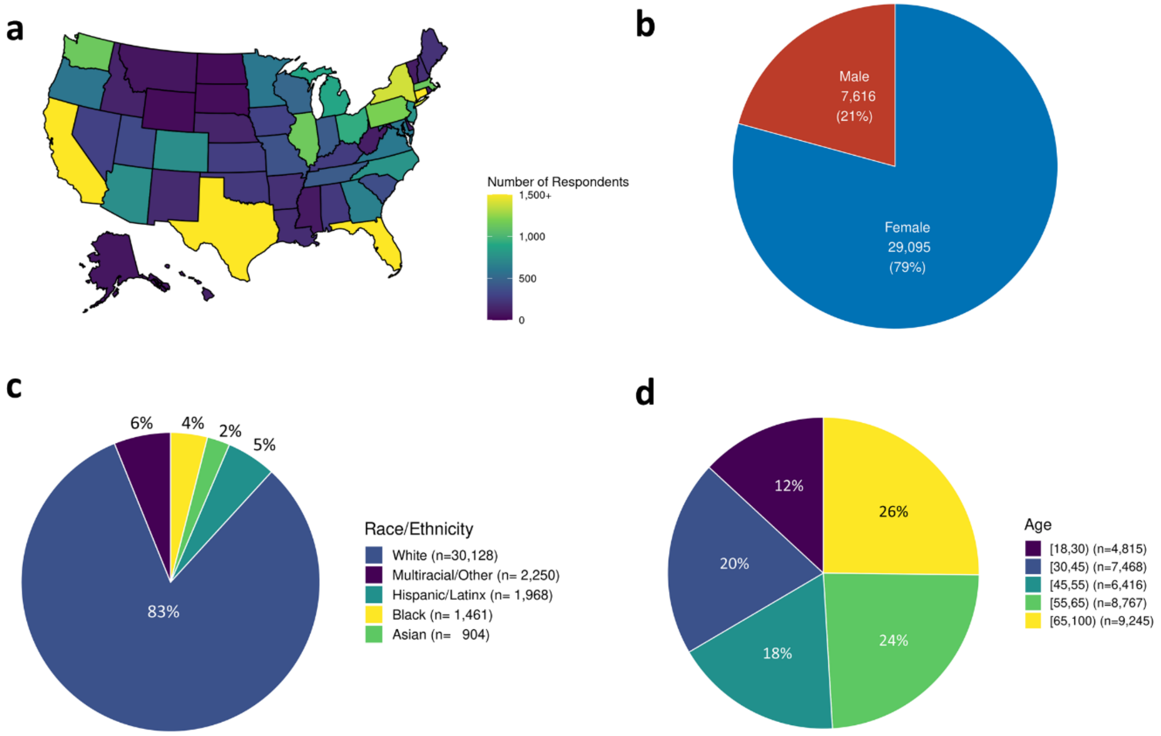


**Supplementary Figure 1, HWF Vaccine Intent Demographic Distributions :** Demographic break down of the HWF user base that responded to the vaccine intent question by **(a)** state, **(b)** sex, **(c)** race and **(d)** age.

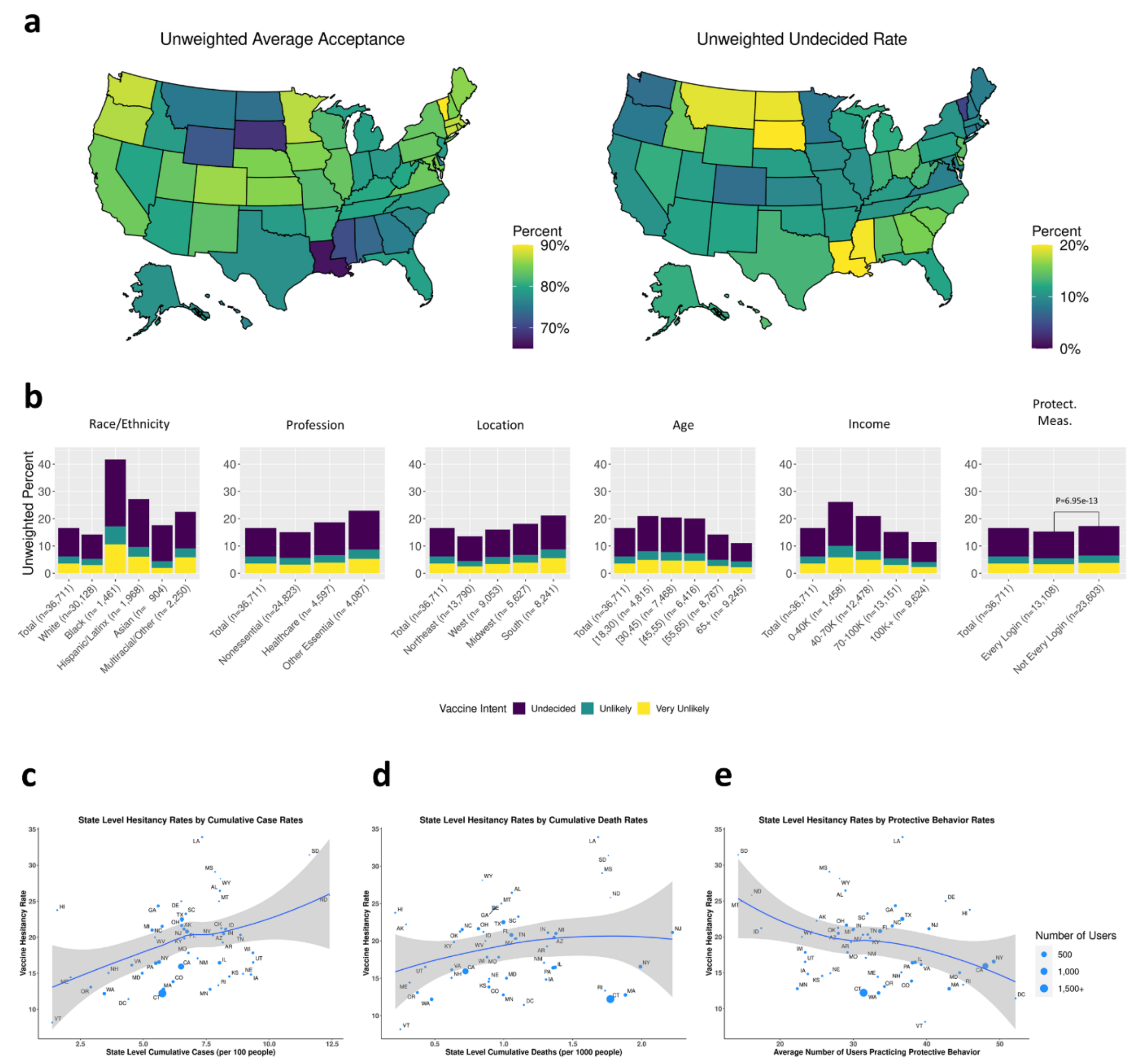


**Supplementary Figure 2, Unweighted vaccine intent responses: (a)** Unweighted statewide (Left) acceptance and (Right) undecided rates. **(b)** Unweighted vaccine responses across several demographic characteristics. State level hesitancy rates estimated using the HowWeFeel data by **(c)** cumulative case (per 100 individuals) and **(d)** death (per 1000 individuals) rates reported in USA Facts and **(e)** average number of users practicing protective behavior.

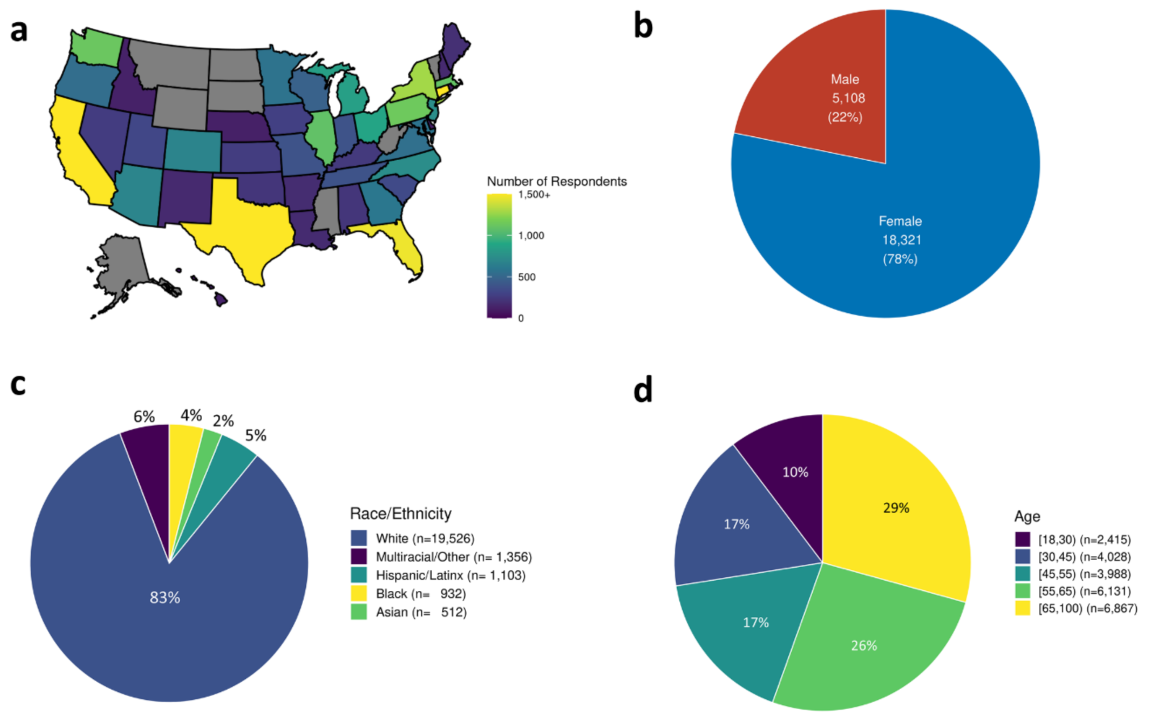


**Supplementary Figure 3, HWF Vaccine Uptake Demographic Distributions :** Demographic break down of the HWF user base that responded to the vaccine uptake question by **(a)** state, **(b)** sex, **(c)** race and **(d)** age.

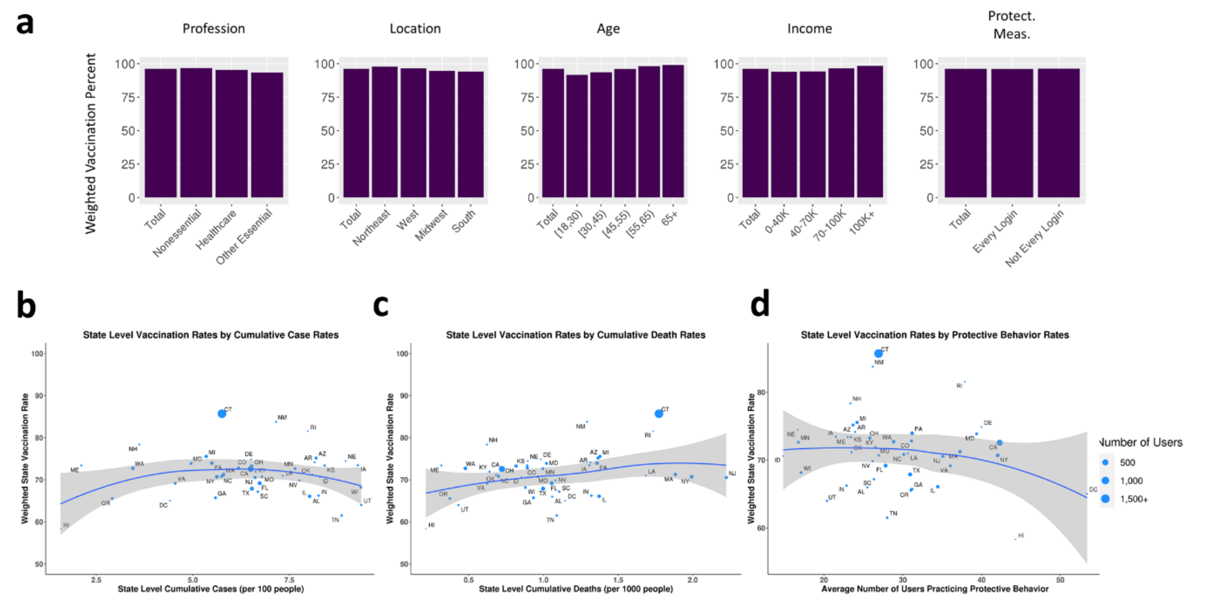


**Supplementary Figure 4, Demographic specific weighted vaccine uptake rates: (a)** Weighted vaccination rates of users that were offered a vaccine by profession, location, age, income, and use of protective measures. State level weighted vaccination rates of users that were offered a vaccine by **(c)** cumulative case rates (/100 individuals), **(d)** cumulative death rates (/1000 individuals), **(e)** and average number of users practicing protective behavior.

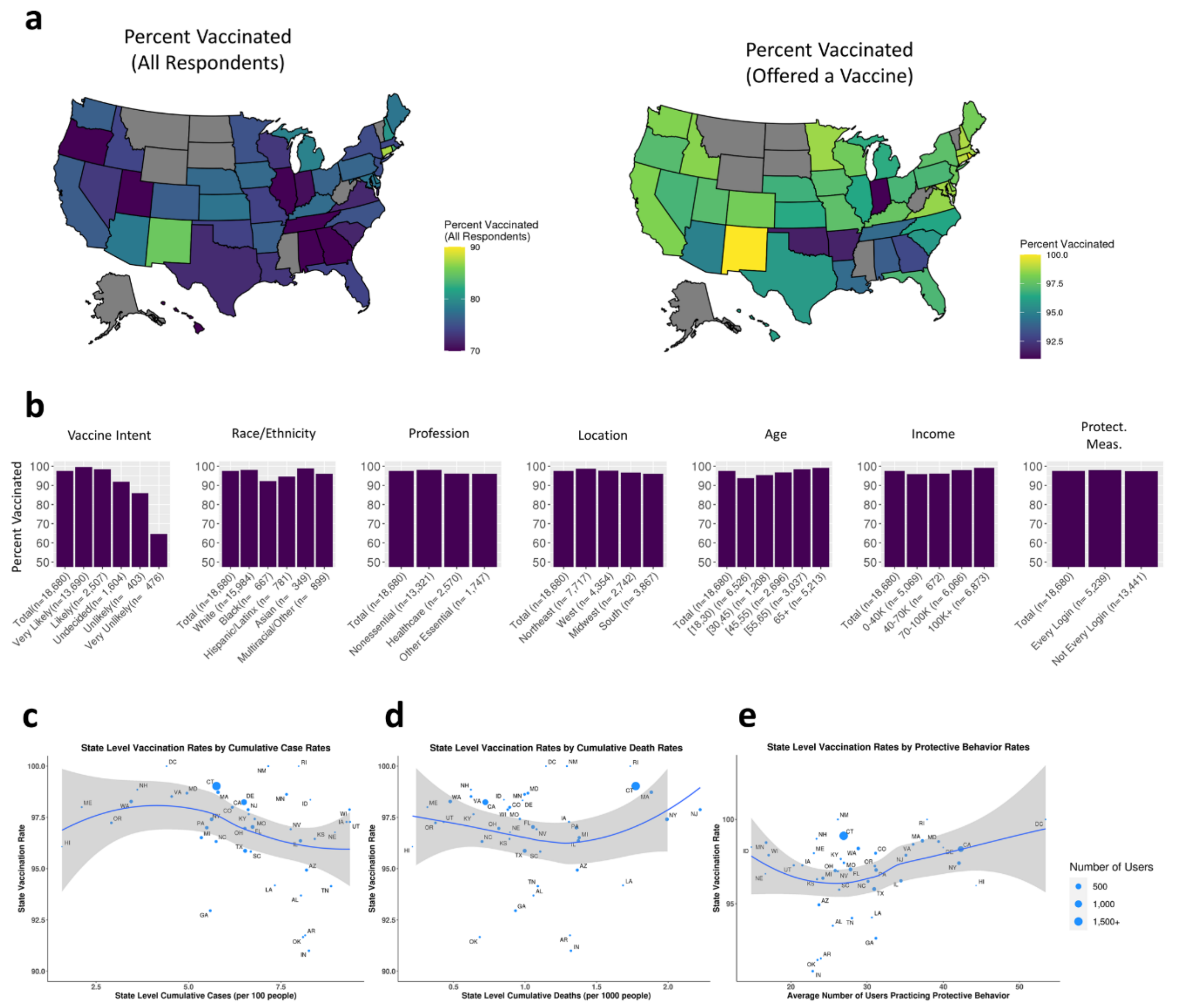


**Supplementary Figure 5, Demographic Specific Vaccine Uptake Rates: (a)** Unweighted vaccination rates by state of (Left) all users that responded to the vaccine uptake question and (right) users that were offered a vaccine. **(b)** Vaccination rates of users that were offered a vaccine by vaccine intent, race/ethnicity, profession, location, age, income, and use of protective measures. **(c)** State level vaccination rates of users that were offered a vaccine by **(b)** cumulative case rates (/100 individuals), **(c)** cumulative death rates (/1000 individuals), **(d)** and average number of users practicing protective behavior.

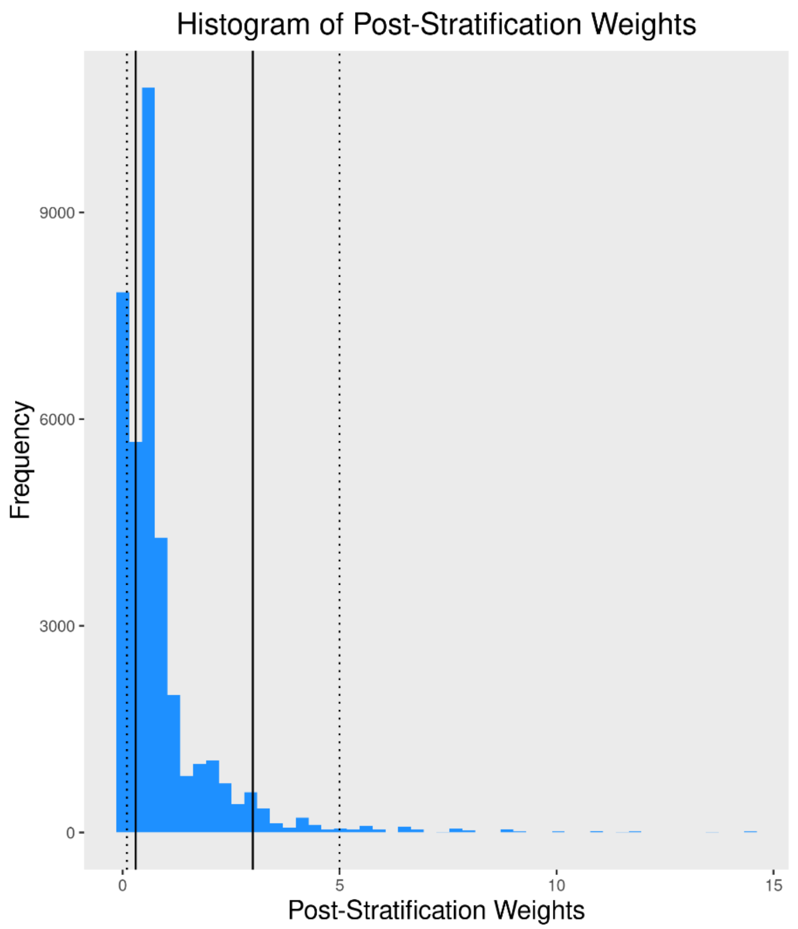


**Supplementary Figure 6, Post-Stratification Weights:** Histogram of post-stratification weights adjusted for census location, race, age, and sex. Solid lines indicate the trimming threshold and the dotted line indicates the trimming threshold for the sensitivity analysis.
