## Supplementary Figures for "Unraveling Attributes of COVID-19 Vaccine Hesitancy and Uptake in the U.S.: A Large Nationwide Study"

**Supplementary Table 1: Demographics** – Descriptive statistics for the demographic characteristics of the HWF user base.

| Variable | Level | Intent Respondents (n = 36,711) | Uptake Respondents (n=23,429) |
|---|---|---|---|
| **Sex** | | | |
| | **Male** | 7,616 (21%) | 5,108 (22%) |
| | **Female** | 29,095 (79%) | 18,321 (78%) |
| **Age** | | | |
| | **[18,30)** | 4,815 (13%) | 2,415 (10%) |
| | **[30,45)** | 7,468 (20%) | 4,028 (17%) |
| | **[45,55)** | 6,416 (17%) | 3,988 (17%) |
| | **[55,65)** | 8,767 (24%) | 6,131 (26%) |
| | **[65,100)** | 9,245 (25%) | 6,867 (29%) |
| **Profession** | | | |
| | **Nonessential** | 24,823 (68%) | 16,492 (70%) |
| | **Healthcare** | 4,597 (13%) | 2,773 (12%) |
| | **Other Essential** | 4,087 (11%) | 2,529 (11%) |
| | **Missing** | 3,204 (9%) | 1,635 (7%) |
| **Employment Status** | | | |
| | **Employed** | 19,205 (52%) | 12,206 (52%) |
| | **Furloughed/Job Seeker** | 2,344 (6%) | 1,466 (6%) |
| | **Unemployed** | 10,264 (28%) | 7,581 (32%) |
| | **Missing** | 4,898 (13%) | 2,176 (9%) |
| **Race/Ethnicity** | | | |
| | **White** | 30,128 (83%) | 19,526 (83%) |
| | **Black/African American** | 1,461 (4%) | 932 (4%) |
| | **Hispanic/Latinx** | 1,968 (5%) | 1,103 (5%) |
| | **Asian** | 904 (2%) | 512 (2%) |
| | **Multiracial/Other** | 2,250 (6%) | 1,356 (6%) |
| **Median Household Income** | | | |
| | **0-40k** | 1,458 (4%) | 897 (4%) |
| | **40-70k** | 12,478 (34%) | 7,770 (33%) |
| | **70-100k** | 13,151 (36%) | 8,536 (36%) |
| | **100K+** | 9,624 (26%) | 6,226 (27%) |
| **Population Density** | | | |
| | **0-149 people/sq. mi** | 4,079 (11%) | 2,509 (11%) |
| | **150-999 people/sq. mi** | 13,516 (37%) | 8,497 (36%) |
| | **1000+ people/sq. mi** | 19,116 (52%) | 12,423 (53%) |
| **Preexisting Conditions** | | | |
| | **0** | 8,535 (23%) | 5,266 (22%) |
| | **1** | 12,564 (34%) | 7,905 (34%) |
| | **2** | 8,502 (23%) | 5,446 (23%) |
| | **3+** | 6,723 (18%) | 4,589 (20%) |
| | **Not Say** | 387 (1%) | 223 (1%) |
| **Census Location** | | | |
| | **Northeast** | 13,790 (38%) | 9,139 (39%) |
| | **West** | 9,053 (25%) | 5,649 (24%) |
| | **Midwest** | 5,627 (15%) | 3,561 (15%) |
| | **South** | 8,241 (22%) | 5,080 (22%) |
| **Practiced Protective Measures** | | | |
| | **During Every Login** | 13,108 (36%) | 7,146 (31%) |
| | **Not Every Login** | 23,603 (64%) | 16,283 (69%) |
| **Ever Received COVID Test** | | | |
| | **Yes** | 5,075 (14%) | 3,262 (14%) |
| | **No** | 31,636 (86%) | 20,167 (86%) |
| **COVID Status** | | | |
| | **Never Tested Positive/Never Tested** | 36,536 (99%) | 107 (<1%) |
| | **Tested Positive** | 175 (<1%) | 23,322 (99%) |

**Supplementary Table 2: Hesitancy Rates** – Unweighted and weighted percentages of the vaccine intent responses across demographics.

| | | Unweighted Percents | | | | | Weighted Percents | | | | |
|---|---|---|---|---|---|---|---|---|---|---|---|
| **Variable** | **Level** | **Very Unlikely (4%)** | **Unlikely (3%)** | **Undecided (10%)** | **Likely (13%)** | **Very Likely (70%)** | **Very Unlikely (4%)** | **Unlikely (3%)** | **Undecided (11%)** | **Likely (13%)** | **Very Likely (69%)** |
| **Sex** | | | | | | | | | | | |
| | **Male** | 188 (2%) | 141 (2%) | 487 (6%) | 765 (10%) | 6035 (79%) | 3% | 2% | 8% | 11% | 76% |
| | **Female** | 1140 (4%) | 802 (3%) | 3335 (11%) | 3980 (14%) | 19838 (68%) | 5% | 3% | 14% | 14% | 64% |
| **Age** | | | | | | | | | | | |
| | **[18,30)** | 237 (5%) | 152 (3%) | 620 (13%) | 747 (16%) | 3059 (64%) | 6% | 3% | 14% | 15% | 62% |
| | **[30,45)** | 352 (5%) | 227 (3%) | 947 (13%) | 1028 (14%) | 4914 (66%) | 5% | 3% | 13% | 13% | 65% |
| | **[45,55)** | 296 (5%) | 175 (3%) | 821 (13%) | 866 (13%) | 4258 (66%) | 5% | 3% | 13% | 13% | 66% |
| | **[55,65)** | 236 (3%) | 196 (2%) | 810 (9%) | 1065 (12%) | 6460 (74%) | 3% | 2% | 10% | 12% | 73% |
| | **[65,100)** | 207 (2%) | 193 (2%) | 624 (7%) | 1039 (11%) | 7182 (78%) | 3% | 2% | 7% | 11% | 77% |
| **Profession** | | | | | | | | | | | |
| | **Nonessential** | 793 (3%) | 603 (2%) | 2345 (9%) | 3241 (13%) | 17841 (72%) | 4% | 3% | 10% | 13% | 70% |
| | **Healthcare** | 177 (4%) | 130 (3%) | 551 (12%) | 548 (12%) | 3191 (69%) | 4% | 3% | 13% | 12% | 68% |
| | **Other Essential** | 217 (5%) | 139 (3%) | 581 (14%) | 580 (14%) | 2570 (63%) | 6% | 4% | 14% | 14% | 62% |
| | **Missing** | 141 (4%) | 71 (2%) | 345 (11%) | 376 (12%) | 2271 (71%) | 5% | 2% | 12% | 12% | 69% |
| **Self-Reported Race/Ethnicity** | | | | | | | | | | | |
| | **White** | 904 (3%) | 682 (2%) | 2696 (9%) | 3755 (12%) | 22091 (73%) | 3% | 2% | 8% | 12% | 74% |
| | **Black/African American** | 155 (11%) | 96 (7%) | 358 (25%) | 235 (16%) | 617 (42%) | 11% | 7% | 25% | 16% | 42% |
| | **Hispanic/Latinx** | 120 (6%) | 70 (4%) | 346 (18%) | 258 (13%) | 1174 (60%) | 6% | 3% | 17% | 13% | 60% |
| | **Asian** | 18 (2%) | 22 (2%) | 119 (13%) | 151 (17%) | 594 (66%) | 2% | 3% | 14% | 15% | 66% |
| | **Multiracial/Other** | 131 (6%) | 73 (3%) | 303 (13%) | 346 (15%) | 1397 (62%) | 6% | 3% | 13% | 14% | 65% |
| **Median Household Income** | | | | | | | | | | | |
| | **0-40k** | 85 (6%) | 62 (4%) | 234 (16%) | 170 (12%) | 907 (62%) | 6% | 5% | 17% | 11% | 61% |
| | **40-70k** | 618 (5%) | 393 (3%) | 1605 (13%) | 1645 (13%) | 8217 (66%) | 6% | 3% | 14% | 13% | 65% |
| | **70-100k** | 402 (3%) | 312 (2%) | 1281 (10%) | 1715 (13%) | 9441 (72%) | 4% | 2% | 10% | 13% | 71% |
| | **100K+** | 223 (2%) | 176 (2%) | 702 (7%) | 1215 (13%) | 7308 (76%) | 3% | 2% | 8% | 13% | 75% |

| | | | | | | | | | | | |
|---|---|---|---|---|---|---|---|---|---|---|---|
| **Population Density** | | | | | | | | | | | |
| | **0-149 people/sq. mi** | 199 (5%) | 131 (3%) | 542 (13%) | 575 (14%) | 2632 (65%) | 5% | 3% | 14% | 14% | 65% |
| | **150-999 people/sq. mi** | 579 (4%) | 379 (3%) | 1448 (11%) | 1726 (13%) | 9384 (69%) | 5% | 3% | 11% | 12% | 68% |
| | **1000+ people/sq. mi** | 550 (3%) | 433 (2%) | 1832 (10%) | 2444 (13%) | 13857 (72%) | 4% | 3% | 11% | 13% | 70% |
| **Preexisting Conditions** | | | | | | | | | | | |
| | **0** | 295 (3%) | 197 (2%) | 869 (10%) | 1121 (13%) | 6053 (71%) | 4% | 3% | 11% | 13% | 69% |
| | **1** | 412 (3%) | 343 (3%) | 1336 (11%) | 1618 (13%) | 8855 (70%) | 4% | 3% | 12% | 13% | 68% |
| | **2** | 302 (4%) | 231 (3%) | 840 (10%) | 1119 (13%) | 6010 (71%) | 4% | 3% | 11% | 12% | 70% |
| | **3+** | 293 (4%) | 164 (2%) | 734 (11%) | 838 (12%) | 4694 (70%) | 5% | 3% | 12% | 12% | 69% |
| | **Not Say** | 26 (7%) | 8 (2%) | 43 (11%) | 49 (13%) | 261 (67%) | 9% | 2% | 14% | 14% | 61% |
| **Census Location** | | | | | | | | | | | |
| | **Northeast** | 345 (3%) | 277 (2%) | 1255 (9%) | 1780 (13%) | 10133 (73%) | 3% | 2% | 10% | 13% | 71% |
| | **West** | 307 (3%) | 240 (3%) | 902 (10%) | 1174 (13%) | 6430 (71%) | 3% | 3% | 10% | 13% | 71% |
| | **Midwest** | 216 (4%) | 164 (3%) | 641 (11%) | 715 (13%) | 3891 (69%) | 4% | 3% | 11% | 13% | 70% |
| | **South** | 460 (6%) | 262 (3%) | 1024 (12%) | 1076 (13%) | 5419 (66%) | 6% | 3% | 13% | 13% | 65% |
| **Employment Status** | | | | | | | | | | | |
| | **Employed** | 661 (3%) | 512 (3%) | 2108 (11%) | 2619 (14%) | 13305 (69%) | 4% | 3% | 11% | 14% | 69% |
| | **Furloughed/Job Seeker** | 102 (4%) | 90 (4%) | 303 (13%) | 310 (13%) | 1539 (66%) | 5% | 4% | 16% | 12% | 63% |
| | **Unemployed** | 320 (3%) | 227 (2%) | 808 (8%) | 1287 (13%) | 7622 (74%) | 4% | 2% | 9% | 12% | 72% |
| | **Missing** | 245 (5%) | 114 (2%) | 603 (12%) | 529 (11%) | 3407 (70%) | 6% | 3% | 13% | 11% | 67% |
| **Practiced Protective Measures** | | | | | | | | | | | |
| | **During Every Login** | 438 (3%) | 287 (2%) | 1282 (10%) | 1475 (11%) | 9626 (73%) | 4% | 2% | 11% | 11% | 72% |
| | **Not Every Login** | 890 (4%) | 656 (3%) | 2540 (11%) | 3270 (14%) | 16247 (69%) | 4% | 3% | 12% | 14% | 67% |
| **Ever Received COVID Test** | | | | | | | | | | | |
| | **Yes** | 145 (3%) | 100 (2%) | 462 (9%) | 569 (11%) | 3799 (75%) | 3% | 2% | 10% | 11% | 74% |
| | **No** | 1183 (4%) | 843 (3%) | 3360 (11%) | 4176 (13%) | 22074 (70%) | 4% | 3% | 12% | 13% | 68% |

**Supplementary Table 3: Univariate Analysis** – Unweighted and weighted univariate logistic regression analyses for vaccine hesitancy.

| Variable | Reference Group | Univariate Analysis | | | | | Weighted Univariate Analysis | | | | |
|---|---|---|---|---|---|---|---|---|---|---|---|
| | | OR | Lower 95% CI | Upper 95% CI | P-Value | Sig | OR | Lower 95% CI | Upper 95% CI | P-Value | Sig |
| Female | Male | 1.85 | 1.71 | 2.00 | 6.49E-53 | *** | 1.91 | 1.74 | 2.09 | 2.54E-43 | *** |
| Age [18,30) | 65+ | 2.13 | 1.94 | 2.34 | 1.07E-54 | *** | 2.10 | 1.87 | 2.36 | 4.58E-35 | *** |
| Age [30,45) | | 2.06 | 1.89 | 2.25 | 3.33E-61 | *** | 1.93 | 1.73 | 2.15 | 4.81E-32 | *** |
| Age [45,55) | | 2.02 | 1.85 | 2.21 | 2.94E-54 | *** | 1.90 | 1.69 | 2.13 | 1.13E-27 | *** |
| Age [55,65) | | 1.33 | 1.21 | 1.45 | 4.44E-10 | *** | 1.24 | 1.11 | 1.39 | 0.0002 | ** |
| Essential Healthcare | Nonessential | 1.29 | 1.19 | 1.40 | 7.76E-10 | *** | 1.32 | 1.19 | 1.46 | 1.01E-07 | *** |
| Other Essential | | 1.68 | 1.55 | 1.82 | 5.03E-36 | *** | 1.56 | 1.41 | 1.73 | 2.25E-17 | *** |
| Furloughed/Job Seeker | Employed | 1.30 | 1.17 | 1.44 | 1.31E-06 | *** | 1.52 | 1.33 | 1.73 | 6.56E-10 | *** |
| Unemployed | | 0.74 | 0.69 | 0.79 | 3.34E-18 | *** | 0.83 | 0.76 | 0.90 | 2.39E-05 | *** |
| African American/Black | White | 4.31 | 3.87 | 4.81 | 1.52E-152 | *** | 4.52 | 4.01 | 5.10 | 1.14E-131 | *** |
| Hispanic/Latinx | | 2.26 | 2.03 | 2.51 | 7.17E-53 | *** | 2.24 | 2.00 | 2.52 | 2.73E-43 | *** |
| Asian | | 1.29 | 1.08 | 1.53 | 0.0044 | * | 1.46 | 1.21 | 1.77 | 0.0001 | ** |
| Multiracial/Other | | 1.76 | 1.58 | 1.95 | 2.89E-26 | *** | 1.66 | 1.47 | 1.89 | 2.37E-15 | *** |
| 1 Preexisting | 0 | 1.05 | 0.98 | 1.13 | 0.1794 | | 1.05 | 0.95 | 1.15 | 0.3305 | |
| 2 Preexisting | | 1.02 | 0.94 | 1.10 | 0.7181 | | 1.01 | 0.91 | 1.12 | 0.8746 | |
| 3+ Preexisting | | 1.13 | 1.04 | 1.24 | 0.0037 | * | 1.08 | 0.97 | 1.21 | 0.1479 | |
| Preexisting Not Say | | 1.31 | 1.01 | 1.68 | 0.0392 | * | 1.51 | 1.08 | 2.09 | 0.0146 | * |
| Parent | Not a Parent | 1.04 | 0.69 | 1.52 | 0.8458 | | 1.17 | 0.72 | 1.90 | 0.5339 | |
| Income 0-40K | 100K+ | 0.96 | 0.91 | 1.02 | 0.190 | | 0.99 | 0.92 | 1.06 | 0.7220 | |
| Income 40-70K | | 2.74 | 2.40 | 3.12 | 3.95E-50 | *** | 2.69 | 2.29 | 3.16 | 1.50E-33 | *** |
| Income 70-100K | | 2.05 | 1.90 | 2.22 | 1.41E-76 | *** | 2.03 | 1.84 | 2.24 | 8.79E-46 | *** |
| PopDensity 0-149 | 1000+ | 1.38 | 1.28 | 1.50 | 6.02E-16 | *** | 1.36 | 1.22 | 1.50 | 6.30E-09 | *** |
| PopDensity 150-999 | | 1.57 | 1.45 | 1.71 | 1.03E-25 | *** | 1.37 | 1.24 | 1.53 | 2.11E-09 | *** |
| West | Northeast | 1.25 | 1.18 | 1.33 | 9.01E-14 | *** | 1.22 | 1.13 | 1.32 | 2.20E-07 | *** |
| Midwest | | 1.21 | 1.12 | 1.30 | 5.33E-07 | *** | 1.07 | 0.97 | 1.18 | 0.1640 | |
| South | | 1.41 | 1.29 | 1.53 | 1.08E-15 | *** | 1.18 | 1.06 | 1.31 | 0.0026 | * |
| Practiced Protective Behavior | Did Not Practice | 1.06 | 1.05 | 1.07 | 1.32E-38 | *** | 1.04 | 1.03 | 1.05 | 8.19E-15 | *** |

| | | | | | | | | | | | |
|---|---|---|---|---|---|---|---|---|---|---|---|
| **Answered before Pfizer EUA** | **After Pfizer EUA** | 1.15 | 1.09 | 1.21 | 8.79E-07 | *** | 1.15 | 1.07 | 1.23 | 0.0001 | ** |
| **State Cumulative Case Rate** | | 1.71 | 1.59 | 1.83 | 3.99E-48 | *** | 1.52 | 1.39 | 1.67 | 4.50E-19 | *** |
| **State Cumulative Death Rate** | | 1.09 | 1.07 | 1.11 | 5.12E-16 | *** | 1.05 | 1.02 | 1.08 | 8.63E-05 | *** |
| **Average Daily Pos Test Rate (Nov.)** | | 0.85 | 0.81 | 0.90 | 1.64E-08 | *** | 0.96 | 0.90 | 1.04 | 0.3131 | |
| **County % without HS Degree** | | 1.03 | 1.03 | 1.04 | 3.11E-30 | *** | 1.03 | 1.02 | 1.04 | 2.45E-16 | *** |
| **Received COVID Test** | **Never Received Test** | 0.79 | 0.72 | 0.86 | 4.05E-08 | *** | 0.78 | 0.70 | 0.87 | 9.41E-06 | *** |

*: $p < 0.05$
**: $p < 0.001$
***: $p < 0.0001$

**Supplementary Table 4: Multivariable Analysis** – Unweighted and weighted multivariable logistic regression analyses for vaccine hesitancy.

| | | Multivariable Unweighted Analysis | | | | | Multivariable Weighted Analysis | | | | |
|---|---|---|---|---|---|---|---|---|---|---|---|
| **Variable** | **Reference Group** | **OR** | **Lower 95% CI** | **Upper 95% CI** | **P-Value** | **Sig** | **OR** | **Lower 95% CI** | **Upper 95% CI** | **P-Value** | **Sig** |
| **Female** | **Male** | 1.73 | 1.59 | 1.88 | 1.41E-38 | *** | 1.67 | 1.51 | 1.83 | 4.09E-25 | *** |
| **Age [18,30)** | **65+** | 1.96 | 1.73 | 2.21 | 3.27E-27 | *** | 2.17 | 1.86 | 2.53 | 1.03E-22 | *** |
| **Age [30,45)** | | 1.94 | 1.75 | 2.15 | 1.15E-37 | *** | 2.02 | 1.77 | 2.29 | 8.92E-27 | *** |
| **Age [45,55)** | | 1.89 | 1.71 | 2.10 | 9.39E-35 | *** | 1.96 | 1.72 | 2.23 | 1.52E-23 | *** |
| **Age [55,65)** | | 1.26 | 1.15 | 1.39 | 2.11E-06 | *** | 1.27 | 1.12 | 1.44 | 0.0002 | ** |
| **Essential Healthcare** | **Nonessential** | 1.18 | 1.08 | 1.29 | 0.0002 | ** | 1.20 | 1.08 | 1.33 | 0.0010 | * |
| **Other Essential** | | 1.57 | 1.44 | 1.72 | 1.91E-24 | *** | 1.52 | 1.36 | 1.70 | 1.42E-13 | *** |
| **Furloughed/Job Seeker** | **Employed** | 1.30 | 1.16 | 1.45 | 4.40E-06 | *** | 1.48 | 1.29 | 1.70 | 4.04E-08 | *** |
| **Unemployed** | | 0.99 | 0.91 | 1.08 | 0.8267 | | 1.13 | 1.01 | 1.25 | 0.0275 | * |
| **African American/Black** | **White** | 3.84 | 3.42 | 4.31 | 2.80E-114 | *** | 3.94 | 3.47 | 4.48 | 1.26E-96 | *** |
| **Hispanic/Latinx** | | 1.85 | 1.65 | 2.07 | 1.59E-26 | *** | 1.89 | 1.67 | 2.14 | 1.99E-23 | *** |
| **Asian** | | 1.49 | 1.24 | 1.78 | 1.71E-05 | *** | 1.80 | 1.46 | 2.21 | 2.05E-08 | *** |
| **Multiracial/Other** | | 1.55 | 1.39 | 1.73 | 3.10E-15 | *** | 1.59 | 1.39 | 1.81 | 8.11E-12 | *** |
| **1 Preexisting** | **0** | 1.07 | 0.99 | 1.15 | 0.0954 | | 1.06 | 0.96 | 1.17 | 0.2144 | |
| **2 Preexisting** | | 1.04 | 0.95 | 1.13 | 0.4081 | | 1.02 | 0.92 | 1.14 | 0.6578 | |
| **3+ Preexisting** | | 1.25 | 1.14 | 1.37 | 2.47E-06 | *** | 1.19 | 1.06 | 1.34 | 0.0036 | * |
| **Preexisting Not Say** | | 1.39 | 1.05 | 1.81 | 0.0169 | * | 1.58 | 1.09 | 2.30 | 0.0161 | * |
| **Parent** | **Not a Parent** | 1.20 | 1.12 | 1.29 | 2.25E-07 | *** | 1.26 | 1.15 | 1.38 | 9.61E-07 | *** |
| **Income 0-40K** | **100K+** | 1.64 | 1.42 | 1.90 | 4.20E-11 | *** | 1.66 | 1.39 | 1.98 | 2.33E-08 | *** |
| **Income 40-70K** | | 1.56 | 1.43 | 1.70 | 2.69E-24 | *** | 1.58 | 1.42 | 1.77 | 1.85E-16 | *** |
| **Income 70-100K** | | 1.24 | 1.14 | 1.35 | 2.85E-07 | *** | 1.21 | 1.08 | 1.34 | 0.0005 | ** |
| **PopDensity 0-149** | **1000+** | 1.34 | 1.21 | 1.47 | 4.84E-09 | *** | 1.32 | 1.18 | 1.49 | 3.86E-06 | *** |
| **PopDensity 150-999** | | 1.22 | 1.14 | 1.30 | 5.18E-09 | *** | 1.27 | 1.16 | 1.38 | 5.02E-08 | *** |
| **West** | **Northeast** | 1.01 | 0.87 | 1.18 | 0.8896 | | 1.02 | 0.83 | 1.23 | 0.8805 | |
| **Midwest** | | 1.12 | 0.97 | 1.28 | 0.1227 | | 1.11 | 0.93 | 1.32 | 0.2364 | |
| **South** | | 1.28 | 1.12 | 1.47 | 0.0004 | ** | 1.25 | 1.05 | 1.48 | 0.0105 | * |
| **Answered before Pfizer EUA** | **After Pfizer EUA** | 1.46 | 1.37 | 1.55 | 5.32E-33 | *** | 1.48 | 1.37 | 1.60 | 9.96E-23 | *** |
| **Practiced Protective Behavior** | **Did Not Practice** | 0.79 | 0.74 | 0.84 | 6.95E-13 | *** | 0.78 | 0.72 | 0.85 | 6.12E-09 | *** |

| | | | | | | | | | | | |
|---|---|---|---|---|---|---|---|---|---|---|---|
| **State Cumulative Case Rate** | | 1.00 | 0.97 | 1.03 | 0.9449 | | 1.01 | 0.97 | 1.04 | 0.7826 | |
| **State Cumulative Death Rate** | | 1.20 | 1.07 | 1.36 | 0.0030 | * | 1.18 | 1.01 | 1.38 | 0.0351 | * |
| **Average Daily Pos Test Rate (Nov.)** | | 1.01 | 0.99 | 1.02 | 0.4374 | | 1.00 | 0.99 | 1.02 | 0.9654 | |
| **County % without HS Degree** | | 1.02 | 1.01 | 1.02 | 3.21E-05 | *** | 1.01 | 1.00 | 1.02 | 0.0819 | |
| **Received COVID Test** | **Never Received Test** | 0.79 | 0.72 | 0.87 | 3.10E-07 | *** | 0.79 | 0.71 | 0.89 | 5.88E-05 | *** |

*: $p < 0.05$
**: $p < 0.001$
***: $p < 0.0001$

**Supplementary Table 5: Nominal Unweighted** – Unweighted nominal logistic regression analyses comparing “Undecided” vs “Likely” (Left) and “Unlikely” vs “Likely”.

| Variable | Reference Group | Undecided | | | | | Unlikely | | | | |
|---|---|---|---|---|---|---|---|---|---|---|---|
| | | OR | Lower 95% CI | Upper 95% CI | P-Value | Sig | OR | Lower 95% CI | Upper 95% CI | P-Value | Sig |
| **Female** | **Male** | 1.88 | 1.70 | 2.08 | 2.68E-33 | *** | 1.51 | 1.33 | 1.71 | 1.18E-10 | *** |
| **Age [18,30)** | **65+** | 1.91 | 1.65 | 2.22 | 1.29E-17 | *** | 2.04 | 1.69 | 2.45 | 4.23E-14 | *** |
| **Age [30,45)** | | 1.92 | 1.69 | 2.17 | 7.23E-25 | *** | 1.98 | 1.70 | 2.31 | 3.37E-18 | *** |
| **Age [45,55)** | | 1.92 | 1.70 | 2.18 | 4.64E-25 | *** | 1.84 | 1.58 | 2.15 | 1.39E-14 | *** |
| **Age [55,65)** | | 1.31 | 1.17 | 1.48 | 4.64E-06 | *** | 1.17 | 1.01 | 1.36 | 0.0387 | * |
| **Essential Healthcare** | **Nonessential** | 1.18 | 1.06 | 1.31 | 0.0022 | * | 1.18 | 1.03 | 1.35 | 0.0152 | * |
| **Other Essential** | | 1.56 | 1.41 | 1.73 | 7.61E-17 | *** | 1.60 | 1.40 | 1.82 | 1.42E-12 | *** |
| **Furloughed/Job Seeker** | **Employed** | 1.25 | 1.09 | 1.43 | 0.0013 | * | 1.39 | 1.18 | 1.64 | 9.21E-05 | *** |
| **Unemployed** | | 0.92 | 0.83 | 1.02 | 0.1067 | | 1.12 | 0.99 | 1.27 | 0.0769 | * |
| **African American/Black** | **White** | 3.63 | 3.17 | 4.17 | 1.33E-75 | *** | 4.19 | 3.57 | 4.91 | 5.62E-70 | *** |
| **Hispanic/Latinx** | | 1.95 | 1.71 | 2.23 | 1.25E-22 | *** | 1.69 | 1.43 | 2.01 | 1.96E-09 | *** |
| **Asian** | | 1.77 | 1.44 | 2.18 | 5.61E-08 | *** | 1.01 | 0.73 | 1.41 | 0.9298 | |
| **Multiracial/Other** | | 1.51 | 1.32 | 1.72 | 2.02E-09 | *** | 1.63 | 1.39 | 1.91 | 2.33E-09 | *** |
| **1 Preexisting** | **0** | 1.07 | 0.97 | 1.17 | 0.1557 | | 1.07 | 0.95 | 1.20 | 0.3002 | |
| **2 Preexisting** | | 1.00 | 0.90 | 1.11 | 0.9525 | | 1.11 | 0.97 | 1.26 | 0.1259 | |
| **3+ Preexisting** | | 1.21 | 1.08 | 1.35 | 0.0007 | ** | 1.31 | 1.14 | 1.51 | 0.0001 | ** |
| **Preexisting Not Say** | | 1.21 | 0.86 | 1.70 | 0.2642 | | 1.71 | 1.17 | 2.49 | 0.0058 | * |
| **Parent** | **Not a Parent** | 1.17 | 1.07 | 1.27 | 0.0003 | ** | 1.27 | 1.14 | 1.41 | 1.77E-05 | *** |
| **Income 0-40K** | **100K+** | 1.69 | 1.42 | 2.02 | 5.78E-09 | *** | 1.56 | 1.26 | 1.94 | 6.17E-05 | *** |
| **Income 40-70K** | | 1.57 | 1.42 | 1.75 | 1.86E-17 | *** | 1.54 | 1.35 | 1.76 | 1.81E-10 | *** |
| **Income 70-100K** | | 1.28 | 1.16 | 1.41 | 1.56E-06 | *** | 1.18 | 1.04 | 1.34 | 0.0126 | * |
| **PopDensity 0-149** | **1000+** | 1.31 | 1.17 | 1.48 | 5.08E-06 | *** | 1.38 | 1.19 | 1.60 | 1.61E-05 | *** |
| **PopDensity 150-999** | | 1.13 | 1.05 | 1.23 | 0.0022 | * | 1.38 | 1.24 | 1.52 | 6.71E-10 | *** |
| **West** | **Northeast** | 0.95 | 0.79 | 1.14 | 0.5796 | | 1.14 | 0.90 | 1.44 | 0.2846 | |
| **Midwest** | | 1.03 | 0.87 | 1.22 | 0.7064 | | 1.28 | 1.04 | 1.59 | 0.0214 | * |
| **South** | | 1.13 | 0.96 | 1.34 | 0.1353 | | 1.58 | 1.28 | 1.94 | 1.58E-05 | *** |
| **Answered before Pfizer EUA** | **After Pfizer EUA** | 1.59 | 1.47 | 1.71 | 5.40E-34 | *** | 1.26 | 1.15 | 1.38 | 1.52E-06 | *** |
| **Practiced Protective Behavior** | **Did Not Practice** | 0.82 | 0.76 | 0.89 | 8.19E-07 | *** | 0.73 | 0.66 | 0.81 | 1.59E-09 | *** |

| | | | | | | | | | | | |
|---|---|---|---|---|---|---|---|---|---|---|---|
| **State Cumulative Case Rate** | | 1.01 | 0.97 | 1.05 | 0.7091 | | 0.99 | 0.95 | 1.04 | 0.7155 | |
| **State Cumulative Death Rate** | | 1.21 | 1.04 | 1.41 | 0.0116 | * | 1.19 | 0.99 | 1.44 | 0.0638 | |
| **Average Daily Pos Test Rate (Nov.)** | | 1.01 | 0.99 | 1.03 | 0.1945 | | 1.00 | 0.98 | 1.02 | 0.6997 | |
| **County % without HS Degree** | | 1.01 | 1.00 | 1.02 | 0.0152 | * | 1.02 | 1.01 | 1.03 | 3.29E-05 | *** |
| **Received COVID Test** | **Never Received Test** | 0.82 | 0.74 | 0.92 | 0.0004 | ** | 0.74 | 0.64 | 0.85 | 2.44E-05 | *** |

*: $p < 0.05$
**: $p < 0.001$
***: $p < 0.0001$

**Supplementary Table 6: Nominal Weighted** –
Weighted nominal logistic regression analyses.

| Variable | Reference Group | Undecided Weighted | | | | | Unlikely Weighted | | | | |
|---|---|---|---|---|---|---|---|---|---|---|---|
| | | OR | Lower 95% CI | Upper 95% CI | P-Value | Sig | OR | Lower 95% CI | Upper 95% CI | P-Value | Sig |
| **Female** | **Male** | 1.80 | 1.59 | 2.02 | 1.21E-21 | *** | 1.51 | 1.29 | 1.72 | 1.05E-07 | *** |
| **Age [18,30)** | **65+** | 1.94 | 1.61 | 2.35 | 6.09E-12 | *** | 2.04 | 2.01 | 3.24 | 1.40E-14 | *** |
| **Age [30,45)** | | 1.94 | 1.65 | 2.27 | 3.08E-16 | *** | 1.98 | 1.78 | 2.63 | 7.86E-15 | *** |
| **Age [45,55)** | | 1.98 | 1.68 | 2.32 | 2.23E-16 | *** | 1.84 | 1.58 | 2.36 | 1.43E-10 | *** |
| **Age [55,65)** | | 1.31 | 1.13 | 1.53 | 0.0005 | ** | 1.17 | 0.97 | 1.43 | 0.0954 | |
| **Essential Healthcare** | **Nonessential** | 1.22 | 1.07 | 1.39 | 0.0028 | * | 1.18 | 0.98 | 1.37 | 0.0813 | |
| **Other Essential** | | 1.49 | 1.30 | 1.71 | 7.14E-09 | *** | 1.60 | 1.32 | 1.83 | 1.70E-07 | *** |
| **Furloughed/Job Seeker** | **Employed** | 1.50 | 1.27 | 1.78 | 2.61E-06 | *** | 1.39 | 1.18 | 1.78 | 0.0004 | ** |
| **Unemployed** | | 1.07 | 0.94 | 1.22 | 0.3332 | | 1.12 | 1.04 | 1.44 | 0.0133 | * |
| **African American/Black** | **White** | 3.90 | 3.34 | 4.56 | 1.39E-66 | *** | 4.19 | 3.38 | 4.79 | 1.65E-54 | *** |
| **Hispanic/Latinx** | | 2.04 | 1.76 | 2.37 | 6.98E-21 | *** | 1.69 | 1.39 | 2.03 | 8.68E-08 | *** |
| **Asian** | | 2.12 | 1.68 | 2.68 | 2.56E-10 | *** | 1.01 | 0.86 | 1.76 | 0.2606 | |
| **Multiracial/Other** | | 1.56 | 1.32 | 1.83 | 7.17E-08 | *** | 1.63 | 1.35 | 1.99 | 8.13E-07 | *** |
| **1 Preexisting** | **0** | 1.08 | 0.96 | 1.22 | 0.1770 | | 1.07 | 0.89 | 1.21 | 0.6269 | |
| **2 Preexisting** | | 0.96 | 0.85 | 1.10 | 0.5888 | | 1.11 | 0.95 | 1.32 | 0.1822 | |
| **3+ Preexisting** | | 1.15 | 1.00 | 1.33 | 0.0535 | | 1.31 | 1.02 | 1.46 | 0.0266 | * |
| **Preexisting Not Say** | | 1.33 | 0.83 | 2.13 | 0.2373 | | 1.71 | 1.12 | 3.12 | 0.0169 | * |
| **Parent** | **Not a Parent** | 1.17 | 1.05 | 1.31 | 0.0052 | * | 1.27 | 1.26 | 1.68 | 2.86E-07 | *** |
| **Income 0-40K** | **100K+** | 1.75 | 1.41 | 2.18 | 3.26E-07 | *** | 1.56 | 1.21 | 2.05 | 0.0008 | ** |
| **Income 40-70K** | | 1.62 | 1.42 | 1.85 | 1.74E-12 | *** | 1.54 | 1.31 | 1.83 | 3.75E-07 | *** |
| **Income 70-100K** | | 1.25 | 1.10 | 1.42 | 0.0008 | ** | 1.18 | 0.97 | 1.35 | 0.1152 | |
| **PopDensity 0-149** | **1000+** | 1.31 | 1.14 | 1.51 | 0.0002 | ** | 1.38 | 1.11 | 1.59 | 0.0019 | ** |
| **PopDensity 150-999** | | 1.17 | 1.06 | 1.30 | 0.0025 | * | 1.38 | 1.24 | 1.59 | 1.52E-07 | *** |
| **West** | **Northeast** | 0.96 | 0.75 | 1.22 | 0.7255 | | 1.14 | 0.83 | 1.49 | 0.4735 | |
| **Midwest** | | 1.03 | 0.84 | 1.28 | 0.7637 | | 1.28 | 0.97 | 1.62 | 0.0827 | |
| **South** | | 1.10 | 0.89 | 1.36 | 0.3716 | | 1.58 | 1.19 | 1.97 | 0.0008 | ** |
| **Answered before Pfizer EUA** | **After Pfizer EUA** | 1.60 | 1.45 | 1.76 | 1.51E-21 | *** | 1.26 | 1.18 | 1.50 | 2.68E-06 | *** |
| **Mask Wearing/Protective Measures** | **Did Not Practice** | 0.82 | 0.74 | 0.90 | 8.80E-05 | *** | 0.73 | 0.64 | 0.83 | 1.56E-06 | *** |

| | | | | | | | | | | |
|---|---|---|---|---|---|---|---|---|---|---|
| **State Cumulative Case Rate** | | 1.01 | 0.97 | 1.06 | 0.5969 | | 0.99 | 0.94 | 1.05 | 0.8213 | |
| **State Cumulative Death Rate** | | 1.18 | 0.97 | 1.43 | 0.0919 | | 1.19 | 0.94 | 1.49 | 0.1448 | |
| **Average Daily Pos Test Rate (Nov.)** | | 1.00 | 0.99 | 1.02 | 0.6512 | | 1.00 | 0.97 | 1.02 | 0.5181 | |
| **County % without HS Degree** | | 1.01 | 1.00 | 1.02 | 0.2902 | | 1.02 | 1.00 | 1.02 | 0.1312 | |
| **Received COVID Test** | **Never Received Test** | 0.82 | 0.72 | 0.94 | 0.0050 | * | 0.74 | 0.62 | 0.89 | 0.0013 | * |

*: p < 0.05
**: p < 0.001
***: p < 0.0001

**Supplementary Table 7: Sensitivity Multivariable** – Sensitivity analysis for the multivariable logistic regression analysis for vaccine hesitancy using a new weight threshold of 0.1 to 5.

| | | Multivariable Sensitivity Analysis for Weight Thresholds (0.1,5) | | | | |
|---|---|---|---|---|---|---|
| **Variable** | **Reference Group** | **OR** | **Lower 95% CI** | **Upper 95% CI** | **P-Value** | **Significant** |
| **Female** | **Male** | 1.65 | 1.48 | 1.83 | 5.93E-21 | *** |
| **Age [18,30)** | **65+** | 2.26 | 1.91 | 2.68 | 6.15E-21 | *** |
| **Age [30,45)** | | 2.03 | 1.77 | 2.34 | 4.37E-23 | *** |
| **Age [45,55)** | | 1.98 | 1.71 | 2.28 | 1.05E-20 | *** |
| **Age [55,65)** | | 1.27 | 1.11 | 1.46 | 0.0005 | ** |
| **Essential Healthcare** | **Nonessential** | 1.20 | 1.07 | 1.35 | 0.0022 | * |
| **Other Essential** | | 1.50 | 1.33 | 1.70 | 1.35E-10 | *** |
| **Furloughed/Job Seeker** | **Employed** | 1.48 | 1.27 | 1.72 | 4.37E-07 | *** |
| **Unemployed** | | 1.17 | 1.04 | 1.31 | 0.0097 | * |
| **African American/Black** | **White** | 3.83 | 3.33 | 4.39 | 4.11E-80 | *** |
| **Hispanic/Latinx** | | 1.91 | 1.67 | 2.19 | 7.94E-21 | *** |
| **Asian** | | 1.80 | 1.46 | 2.22 | 3.93E-08 | *** |
| **Multiracial/Other** | | 1.57 | 1.36 | 1.81 | 1.24E-09 | *** |
| **1 Preexisting** | **0** | 1.07 | 0.96 | 1.19 | 0.2529 | |
| **2 Preexisting** | | 1.02 | 0.90 | 1.15 | 0.7673 | |
| **3+ Preexisting** | | 1.19 | 1.05 | 1.36 | 0.0082 | * |
| **Preexisting Not Say** | | 1.61 | 1.06 | 2.43 | 0.0248 | * |
| **Parent** | **Not a Parent** | 1.28 | 1.16 | 1.42 | 2.28E-06 | *** |
| **Income 0-40K** | **100K+** | 1.70 | 1.40 | 2.08 | 1.40E-07 | *** |
| **Income 40-70K** | | 1.59 | 1.41 | 1.80 | 1.02E-13 | *** |
| **Income 70-100K** | | 1.19 | 1.06 | 1.34 | 0.0040 | * |
| **PopDensity 0-149** | **1000+** | 1.30 | 1.14 | 1.48 | 9.26E-05 | *** |
| **PopDensity 150-999** | | 1.29 | 1.17 | 1.42 | 1.90E-07 | *** |
| **West** | **Northeast** | 1.01 | 0.81 | 1.25 | 0.9410 | |
| **Midwest** | | 1.10 | 0.90 | 1.33 | 0.3495 | |
| **South** | | 1.26 | 1.04 | 1.52 | 0.0180 | * |
| **Answered before Pfizer EUA** | **After Pfizer EUA** | 1.47 | 1.35 | 1.61 | 2.73E-18 | *** |
| **Practiced Protective Behavior** | **Did Not Practice** | 0.78 | 0.71 | 0.85 | 1.04E-07 | *** |
| **State Cumulative Case Rate** | | 1.01 | 0.97 | 1.05 | 0.6396 | |
| **State Cumulative Death Rate** | | 1.17 | 0.99 | 1.39 | 0.0650 | |

| | | | | | | |
|---|---|---|---|---|---|---|
| **Average Daily Pos Test Rate (Nov.)** | | 1.00 | 0.98 | 1.01 | 0.7254 | |
| **County % without HS Degree** | | 1.00 | 1.00 | 1.01 | 0.2847 | |
| **Received COVID Test** | **Never Received Test** | 0.78 | 0.68 | 0.88 | 6.52E-05 | *** |

*: $p < 0.05$
**: $p < 0.001$
***: $p < 0.0001$

**Supplementary Table 8: Sensitivity Nominal** - Sensitivity analysis for the nominal logistic regression analysis using a new weight threshold of 0.1 to 5. Left: logistic regression analyses comparing “Undecided” vs “Likely” and right: logistic regression analyses comparing “Unlikely” vs “Likely”.

| | | Undecided Sensitivity Weights (0.1, 5) | | | | | Unlikely Sensitivity Weights (0.1, 5) | | | | |
|---|---|---|---|---|---|---|---|---|---|---|---|
| Variable | Reference Group | OR | Lower 95% CI | Upper 95% CI | P-Value | Sig | OR | Lower 95% CI | Upper 95% CI | P-Value | Sig |
| **Female** | **Male** | 1.79 | 1.57 | 2.03 | 1.08E-18 | *** | 1.46 | 1.25 | 1.71 | 2.57E-06 | *** |
| **Age [18,30)** | **65+** | 1.92 | 1.56 | 2.36 | 5.62E-10 | *** | 2.87 | 2.21 | 3.73 | 3.53E-15 | *** |
| **Age [30,45)** | | 1.91 | 1.61 | 2.28 | 1.79E-13 | *** | 2.24 | 1.82 | 2.77 | 8.28E-14 | *** |
| **Age [45,55)** | | 1.97 | 1.65 | 2.35 | 4.60E-14 | *** | 1.98 | 1.59 | 2.46 | 5.94E-10 | *** |
| **Age [55,65)** | | 1.30 | 1.10 | 1.54 | 0.0019 | * | 1.20 | 0.98 | 1.48 | 0.0848 | |
| **Essential Healthcare** | **Nonessential** | 1.23 | 1.06 | 1.41 | 0.0049 | * | 1.16 | 0.97 | 1.39 | 0.1058 | |
| **Other Essential** | | 1.48 | 1.27 | 1.72 | 3.81E-07 | *** | 1.52 | 1.27 | 1.82 | 6.39E-06 | *** |
| **Furloughed/Job Seeker** | **Employed** | 1.52 | 1.27 | 1.83 | 6.91E-06 | *** | 1.42 | 1.13 | 1.78 | 0.0022 | * |
| **Unemployed** | | 1.10 | 0.96 | 1.27 | 0.1748 | | 1.27 | 1.06 | 1.51 | 0.0095 | * |
| **African American/Black** | **White** | 3.87 | 3.28 | 4.57 | 1.92E-57 | *** | 3.77 | 3.13 | 4.56 | 2.57E-43 | *** |
| **Hispanic/Latinx** | | 2.11 | 1.79 | 2.48 | 3.34E-19 | *** | 1.65 | 1.35 | 2.03 | 1.26E-06 | *** |
| **Asian** | | 2.14 | 1.69 | 2.72 | 3.97E-10 | *** | 1.20 | 0.84 | 1.73 | 0.3201 | |
| **Multiracial/Other** | | 1.55 | 1.30 | 1.84 | 8.74E-07 | *** | 1.60 | 1.29 | 1.98 | 2.16E-05 | *** |
| **1 Preexisting** | **0** | 1.09 | 0.96 | 1.25 | 0.1766 | | 1.03 | 0.87 | 1.22 | 0.7387 | |
| **2 Preexisting** | | 0.95 | 0.82 | 1.10 | 0.4932 | | 1.12 | 0.93 | 1.35 | 0.2247 | |
| **3+ Preexisting** | | 1.15 | 0.98 | 1.35 | 0.0768 | | 1.22 | 1.00 | 1.49 | 0.0481 | * |
| **Preexisting Not Say** | | 1.34 | 0.79 | 2.27 | 0.2733 | | 1.90 | 1.09 | 3.32 | 0.0246 | * |
| **Parent** | **Not a Parent** | 1.17 | 1.03 | 1.33 | 0.0127 | * | 1.52 | 1.30 | 1.78 | 2.84E-07 | *** |
| **Income 0-40K** | **100K+** | 1.79 | 1.41 | 2.28 | 2.14E-06 | *** | 1.62 | 1.21 | 2.17 | 0.0013 | * |
| **Income 40-70K** | | 1.62 | 1.39 | 1.88 | 2.75E-10 | *** | 1.56 | 1.30 | 1.88 | 2.51E-06 | *** |
| **Income 70-100K** | | 1.23 | 1.07 | 1.42 | 0.0049 | * | 1.13 | 0.94 | 1.36 | 0.1918 | |
| **PopDensity 0-149** | **1000+** | 1.27 | 1.09 | 1.49 | 0.0022 | * | 1.32 | 1.08 | 1.61 | 0.0071 | * |
| **PopDensity 150-999** | | 1.19 | 1.06 | 1.34 | 0.0039 | * | 1.43 | 1.25 | 1.65 | 4.86E-07 | *** |
| **West** | **Northeast** | 0.92 | 0.70 | 1.20 | 0.5409 | | 1.17 | 0.84 | 1.61 | 0.3498 | |
| **Midwest** | | 0.99 | 0.79 | 1.26 | 0.9603 | | 1.29 | 0.97 | 1.70 | 0.0779 | |
| **South** | | 1.07 | 0.85 | 1.35 | 0.5636 | | 1.62 | 1.23 | 2.14 | 0.0006 | ** |

| | | | | | | | | | | | |
|---|---|---|---|---|---|---|---|---|---|---|---|
| **Answered before Pfizer EUA** | **After Pfizer EUA** | 1.59 | 1.43 | 1.77 | 1.09E-17 | *** | 1.33 | 1.16 | 1.51 | 2.65E-05 | *** |
| **Practiced Protective Behavior** | **Did Not Practice** | 0.81 | 0.73 | 0.91 | 0.0003 | ** | 0.73 | 0.64 | 0.84 | 1.34E-05 | *** |
| **State Cumulative Case Rate** | | 1.02 | 0.97 | 1.07 | 0.5056 | | 1.00 | 0.94 | 1.06 | 0.9572 | |
| **State Cumulative Death Rate** | | 1.14 | 0.93 | 1.41 | 0.2149 | | 1.23 | 0.96 | 1.57 | 0.1046 | |
| **Average Daily Pos Test Rate (Nov.)** | | 1.00 | 0.98 | 1.02 | 0.8418 | | 0.99 | 0.96 | 1.01 | 0.2928 | |
| **County % without HS Degree** | | 1.00 | 0.99 | 1.01 | 0.4943 | | 1.01 | 0.99 | 1.02 | 0.4032 | |
| **Received COVID Test** | **Never Received Test** | 0.81 | 0.70 | 0.94 | 0.0060 | * | 0.72 | 0.60 | 0.88 | 0.0010 | * |

*: $p < 0.05$
**: $p < 0.001$
***: $p < 0.0001$

**Supplementary Table 9: Receiving Test** – Weighted logistic regression analysis for receiving a COVID-19 test.

| Name | Reference Group | Probability of Receiving a COVID Test | | | | |
|---|---|---|---|---|---|---|
| | | OR | Lower 95% CI | Upper 95% CI | P-Value | Sig |
| **Female** | **Male** | 0.92 | 0.84 | 1.01 | 0.0939 | |
| **Age [18,30)** | **65+** | 0.68 | 0.58 | 0.81 | 8.46E-06 | *** |
| **Age [30,45)** | | 0.82 | 0.71 | 0.93 | 0.0032 | * |
| **Age [45,55)** | | 0.91 | 0.79 | 1.04 | 0.1670 | |
| **Age [55,65)** | | 0.91 | 0.81 | 1.03 | 0.1455 | |
| **Essential Healthcare** | **Nonessential** | 2.12 | 1.91 | 2.36 | 9.19E-46 | *** |
| **Other Essential** | | 0.97 | 0.85 | 1.11 | 0.6502 | |
| **Furloughed/Job Seeker** | **Employed** | 0.93 | 0.79 | 1.10 | 0.4023 | |
| **Unemployed** | | 0.84 | 0.75 | 0.94 | 0.0022 | * |
| **African American/Black** | **White** | 0.98 | 0.82 | 1.16 | 0.7966 | |
| **Hispanic/Latinx** | | 0.88 | 0.75 | 1.03 | 0.1138 | |
| **Asian** | | 0.96 | 0.77 | 1.21 | 0.7582 | |
| **Multiracial/Other** | | 0.96 | 0.81 | 1.13 | 0.5965 | |
| **1 Preexisting** | **0** | 1.04 | 0.93 | 1.16 | 0.4715 | |
| **2 Preexisting** | | 1.12 | 1.00 | 1.27 | 0.0597 | |
| **3+ Preexisting** | | 1.48 | 1.31 | 1.68 | 6.57E-10 | *** |
| **Preexisting Not Say** | | 0.71 | 0.39 | 1.28 | 0.2550 | |
| **Parent** | **Not a Parent** | 0.95 | 0.87 | 1.05 | 0.3121 | |
| **Income 0-40K** | **100K+** | 1.06 | 0.86 | 1.32 | 0.5750 | |
| **Income 40-70K** | | 0.99 | 0.89 | 1.11 | 0.9141 | |
| **Income 70-100K** | | 1.02 | 0.92 | 1.13 | 0.7056 | |
| **PopDensity 0-149** | **1000+** | 0.78 | 0.67 | 0.90 | 0.0007 | ** |
| **PopDensity 150-999** | | 0.86 | 0.78 | 0.94 | 0.0012 | * |
| **West** | **Northeast** | 1.26 | 1.01 | 1.57 | 0.0438 | * |
| **Midwest** | | 0.81 | 0.66 | 0.99 | 0.0394 | * |
| **South** | | 1.17 | 0.97 | 1.42 | 0.0982 | |
| **Answered before Pfizer EUA** | **After Pfizer EUA** | 0.99 | 0.91 | 1.08 | 0.9030 | |
| **Practiced Protective Behavior** | **Did Not Practice** | 1.02 | 0.94 | 1.12 | 0.6097 | |

| | | | | | |
|---|---|---|---|---|---|
| **State Cumulative Case Rate** | 1.08 | 1.04 | 1.14 | 0.0005 | ** |
| **State Cumulative Death Rate** | 1.40 | 1.18 | 1.68 | 0.0002 | ** |
| **Average Daily Pos Test Rate (Nov.)** | 0.95 | 0.94 | 0.97 | 4.12E-06 | *** |
| **County % without HS Degree** | 1.01 | 1.00 | 1.02 | 0.0240 | * |

*: p < 0.05
**: p < 0.001
***: p < 0.0001

**Supplementary Table 10: IPW Analysis** – IPW analysis utilizing the probability of receiving a COVID-19 test as an inverse probability weight.

| Variable | Reference Group | IPW Trim (0.5,0.95) n=4,837 | | | | | IPW Trim (0.1,0.9) n=4,837 | | | | |
|---|---|---|---|---|---|---|---|---|---|---|---|
| | | OR | Lower 95% CI | Upper 95% CI | P-Value | Sig | OR | Lower 95% CI | Upper 95% CI | P-Value | Sig |
| Female | Male | 1.62 | 1.24 | 2.10 | 0.0003 | ** | 1.62 | 1.25 | 2.10 | 0.0003 | ** |
| Age [18,30) | 65+ | 2.64 | 1.72 | 4.05 | 8.47E-06 | *** | 2.68 | 1.77 | 4.07 | 3.32E-06 | *** |
| Age [30,45) | | 2.03 | 1.46 | 2.84 | 3.24E-05 | *** | 2.01 | 1.45 | 2.79 | 2.86E-05 | *** |
| Age [45,55) | | 2.07 | 1.50 | 2.87 | 1.11E-05 | *** | 2.04 | 1.49 | 2.81 | 1.07E-05 | *** |
| Age [55,65) | | 1.42 | 1.06 | 1.90 | 0.0193 | * | 1.42 | 1.06 | 1.89 | 0.0175 | * |
| Essential Healthcare | Nonessential | 1.28 | 1.04 | 1.59 | 0.0201 | * | 1.28 | 1.04 | 1.58 | 0.0207 | * |
| Other Essential | | 1.16 | 0.87 | 1.53 | 0.3193 | | 1.14 | 0.87 | 1.51 | 0.3434 | |
| Furloughed/Job Seeker | Employed | 1.20 | 0.85 | 1.70 | 0.2991 | | 1.23 | 0.88 | 1.74 | 0.2279 | |
| Unemployed | | 0.97 | 0.74 | 1.27 | 0.8379 | | 0.98 | 0.75 | 1.27 | 0.8633 | |
| African American/Black | White | 4.01 | 2.87 | 5.61 | 6.26E-16 | *** | 3.98 | 2.85 | 5.55 | 5.20E-16 | *** |
| Hispanic/Latinx | | 1.83 | 1.28 | 2.63 | 0.0009 | ** | 1.79 | 1.26 | 2.53 | 0.0010 | * |
| Asian | | 1.78 | 1.01 | 3.16 | 0.0474 | * | 1.76 | 1.00 | 3.12 | 0.0510 | |
| Multiracial/Other | | 1.66 | 1.18 | 2.33 | 0.0037 | * | 1.62 | 1.16 | 2.28 | 0.0051 | * |
| 1 Preexisting | 0 | 1.58 | 1.22 | 2.05 | 0.0006 | ** | 1.59 | 1.23 | 2.04 | 0.0003 | ** |
| 2 Preexisting | | 1.23 | 0.93 | 1.63 | 0.1503 | | 1.26 | 0.96 | 1.66 | 0.0970 | |
| 3+ Preexisting | | 1.63 | 1.22 | 2.17 | 0.0009 | ** | 1.64 | 1.24 | 2.17 | 0.0005 | ** |
| Preexisting Not Say | | 0.73 | 0.15 | 3.51 | 0.6933 | | 0.79 | 0.17 | 3.72 | 0.7646 | |
| Parent | Not a Parent | 1.41 | 1.11 | 1.78 | 0.0044 | * | 1.40 | 1.12 | 1.76 | 0.0037 | * |
| Income 0-40K | 100K+ | 1.65 | 1.03 | 2.63 | 0.0365 | * | 1.66 | 1.05 | 2.63 | 0.0306 | * |
| Income 40-70K | | 1.77 | 1.34 | 2.32 | 4.70E-05 | *** | 1.75 | 1.34 | 2.29 | 4.75E-05 | *** |
| Income 70-100K | | 1.36 | 1.05 | 1.76 | 0.0191 | * | 1.34 | 1.04 | 1.72 | 0.0249 | * |
| PopDensity 0-149 | 1000+ | 1.03 | 0.75 | 1.43 | 0.8506 | | 1.03 | 0.75 | 1.42 | 0.8361 | |
| PopDensity 150-999 | | 1.18 | 0.96 | 1.46 | 0.1254 | | 1.20 | 0.98 | 1.48 | 0.0810 | |
| West | Northeast | 0.65 | 0.39 | 1.09 | 0.1024 | | 0.69 | 0.42 | 1.13 | 0.1373 | |
| Midwest | | 0.94 | 0.59 | 1.49 | 0.7909 | | 1.01 | 0.64 | 1.58 | 0.9771 | |
| South | | 1.21 | 0.77 | 1.88 | 0.4078 | | 1.23 | 0.80 | 1.89 | 0.3354 | |

| | | | | | | | | | | | |
|---|---|---|---|---|---|---|---|---|---|---|---|
| **Answered before Pfizer EUA** | **After Pfizer EUA** | 1.78 | 1.47 | 2.14 | 2.64E-09 | *** | 1.78 | 1.48 | 2.14 | 1.36E-09 | *** |
| **Practiced Protective Behavior** | **Did Not Practice** | 0.92 | 0.76 | 1.12 | 0.4286 | | 0.93 | 0.77 | 1.13 | 0.4940 | |
| **State Cumulative Case Rate** | | 1.01 | 0.91 | 1.12 | 0.8462 | | 1.01 | 0.91 | 1.11 | 0.9124 | |
| **State Cumulative Death Rate** | | 0.83 | 0.56 | 1.23 | 0.3537 | | 0.85 | 0.58 | 1.25 | 0.4028 | |
| **Average Daily Pos Test Rate (Nov.)** | | 0.97 | 0.93 | 1.02 | 0.2570 | | 0.97 | 0.93 | 1.02 | 0.2324 | |
| **County % without HS Degree** | | 1.02 | 1.00 | 1.04 | 0.0633 | | 1.02 | 1.00 | 1.04 | 0.0630 | |
| **Tested Positive** | **Tested Negative** | 1.25 | 0.79 | 1.96 | 0.3443 | | 1.26 | 0.80 | 1.97 | 0.3154 | |

*: p < 0.05
**: p < 0.001
***: p < 0.0001

**Supplementary Table 11: Interim uptake analyses -** Interim analyses for the weighting procedure of the vaccine uptake analysis. Left is the weighted model to predict if a user responded to the vacccine uptake question, while right is the weighted model for predicting if a user was offered a vaccine.

| | | Responded to Vaccine Uptake Question (n = 23,782) | | | | | Offered Vaccine (n = 18,928) | | | | |
|---|---|---|---|---|---|---|---|---|---|---|---|
| **Variable** | **Reference Group** | **OR** | **Lower 95% CI** | **Upper 95% CI** | **P-Value** | **Sig** | **OR** | **Lower 95% CI** | **Upper 95% CI** | **P-Value** | **Sig** |
| **Female** | **Male** | 0.92 | 0.86 | 0.98 | 0.0142 | * | 1.09 | 1.00 | 1.20 | 0.0615 | |
| **Age [18,30)** | **65+** | 0.35 | 0.31 | 0.40 | 4.06E-69 | *** | 0.05 | 0.04 | 0.06 | 5.78E-264 | *** |
| **Age [30,45)** | | 0.45 | 0.41 | 0.50 | 2.90E-57 | *** | 0.09 | 0.08 | 0.11 | 8.67E-215 | *** |
| **Age [45,55)** | | 0.65 | 0.59 | 0.72 | 6.45E-17 | *** | 0.15 | 0.13 | 0.17 | 2.06E-139 | *** |
| **Age [55,65)** | | 0.88 | 0.81 | 0.97 | 0.0079 | * | 0.26 | 0.23 | 0.30 | 3.56E-80 | *** |
| **Essential Healthcare** | **Nonessential** | 0.84 | 0.77 | 0.92 | 0.0002 | ** | 3.90 | 3.30 | 4.60 | 1.06E-57 | *** |
| **Other Essential** | | 0.95 | 0.87 | 1.05 | | | 0.72 | 0.65 | 0.80 | 3.23E-09 | *** |
| **Furloughed/Job Seeker** | **Employed** | 0.93 | 0.83 | 1.05 | 0.2525 | | 0.61 | 0.53 | 0.70 | 1.96E-12 | *** |
| **Unemployed** | | 1.18 | 1.09 | 1.28 | 6.29E-05 | *** | 0.72 | 0.65 | 0.80 | 1.51E-10 | *** |
| **African American/Black** | **White** | 1.32 | 1.16 | 1.50 | 1.85E-05 | *** | 0.96 | 0.80 | 1.14 | 0.6345 | |
| **Hispanic/Latinx** | | 1.00 | 0.89 | 1.12 | 0.9905 | | 1.00 | 0.85 | 1.17 | 0.9727 | |
| **Asian** | | 1.04 | 0.89 | 1.22 | 0.5928 | | 0.98 | 0.77 | 1.24 | 0.8396 | |
| **Multiracial/Other** | | 1.22 | 1.09 | 1.37 | 0.0007 | ** | 0.80 | 0.70 | 0.92 | 0.0016 | * |
| **1 Preexisting** | **0** | 0.97 | 0.90 | 1.05 | 0.4792 | | 1.03 | 0.93 | 1.13 | 0.5779 | |
| **2 Preexisting** | | 0.93 | 0.86 | 1.02 | 0.1189 | | 1.13 | 1.01 | 1.25 | 0.0335 | * |
| **3+ Preexisting** | | 1.04 | 0.95 | 1.15 | 0.3825 | | 1.01 | 0.90 | 1.14 | 0.8321 | |
| **Preexisting Not Say** | | 1.14 | 0.83 | 1.56 | 0.4264 | | 1.06 | 0.74 | 1.51 | 0.7608 | |
| **Parent** | **Not a Parent** | 0.85 | 0.79 | 0.91 | 6.41E-06 | *** | 0.89 | 0.82 | 0.98 | 0.0120 | * |
| **Income 0-40K** | **100K+** | 1.07 | 0.91 | 1.24 | 0.4178 | | 0.98 | 0.80 | 1.20 | 0.8425 | |
| **Income 40-70K** | | 1.06 | 0.97 | 1.15 | 0.2059 | | 0.98 | 0.88 | 1.09 | 0.6805 | |
| **Income 70-100K** | | 1.10 | 1.02 | 1.19 | 0.0179 | * | 0.99 | 0.90 | 1.09 | 0.8639 | |
| **PopDensity 0-149** | **1000+** | 0.83 | 0.75 | 0.92 | 0.0002 | ** | 0.91 | 0.80 | 1.04 | 0.1681 | |
| **PopDensity 150-999** | | 0.85 | 0.80 | 0.91 | 4.20E-06 | *** | 1.02 | 0.93 | 1.11 | 0.6911 | |
| **West** | **Northeast** | 1.11 | 0.95 | 1.30 | 0.1842 | | 0.96 | 0.79 | 1.17 | 0.6803 | |
| **Midwest** | | 1.05 | 0.92 | 1.21 | 0.470 | | 0.91 | 0.77 | 1.09 | 0.3125 | |
| **South** | | 1.02 | 0.89 | 1.17 | 0.7614 | | 0.87 | 0.73 | 1.04 | 0.1201 | |
| **Practiced Protective Behavior** | **Did Not Practice** | 0.62 | 0.58 | 0.66 | 1.35E-51 | *** | 0.66 | 0.61 | 0.72 | 3.38E-24 | *** |

| | | | | | | | | | | | |
|---|---|---|---|---|---|---|---|---|---|---|---|
| **State Cumulative Case Rate** | | 0.98 | 0.95 | 1.01 | 0.2732 | | 1.01 | 0.97 | 1.05 | 0.720 | |
| **State Cumulative Death Rate** | | 1.09 | 0.96 | 1.23 | 0.1729 | | 1.03 | 0.88 | 1.21 | 0.7137 | |
| **Average Daily Pos Test Rate (Nov.)** | | 1.01 | 0.99 | 1.02 | 0.2767 | | 1.00 | 0.98 | 1.01 | 0.6282 | |
| **County % without HS Degree** | | 0.99 | 0.99 | 1.00 | 0.1606 | | 1.01 | 1.00 | 1.02 | 0.1056 | |
| **Received COVID Test** | **Never Received Test** | 0.94 | 0.86 | 1.02 | 0.1488 | | 1.23 | 1.09 | 1.38 | 0.0005 | ** |
| **Vaccine Intent - Unlikely** | **Vaccine Intent - Likely** | 0.84 | 0.75 | 0.94 | 0.0029 | * | 0.51 | 0.44 | 0.59 | 2.53E-19 | *** |
| **Vaccine Intent - Undecided** | **Vaccine Intent - Likely** | 0.91 | 0.83 | 0.99 | 0.0363 | * | 0.57 | 0.51 | 0.64 | 1.33E-22 | *** |

*: $p < 0.05$
**: $p < 0.001$
***: $p < 0.0001$

**Supplementary Table 12: Vaccination Uptake Analyses** - Unweighted model for predicting vaccine uptake (left) and a weighed model for predicting vaccine uptake accounting for non-response bias and biases associated with being offered a vaccine.

| | | Unweighted Uptake Analysis (n = 18,928) | | | | | Weighted Uptake Analysis (n = 18,928) | | | | |
|---|---|---|---|---|---|---|---|---|---|---|---|
| **Variable** | **Reference Group** | **OR** | **Lower 95% CI** | **Upper 95% CI** | **P-Value** | **Sig** | **OR** | **Lower 95% CI** | **Upper 95% CI** | **P-Value** | **Sig** |
| **Female** | **Male** | 0.74 | 0.52 | 1.03 | 0.0808 | | 0.90 | 0.60 | 1.33 | 0.5868 | |
| **Age [18,30)** | **65+** | 0.13 | 0.08 | 0.20 | 2.41E-18 | *** | 0.10 | 0.06 | 0.18 | 1.43E-16 | *** |
| **Age [30,45)** | | 0.23 | 0.16 | 0.34 | 7.06E-14 | *** | 0.20 | 0.13 | 0.31 | 1.29E-13 | *** |
| **Age [45,55)** | | 0.40 | 0.27 | 0.59 | 3.94E-06 | *** | 0.38 | 0.25 | 0.57 | 3.68E-06 | *** |
| **Age [55,65)** | | 0.57 | 0.39 | 0.82 | 0.0025 | * | 0.55 | 0.37 | 0.81 | 0.0029 | * |
| **Essential Healthcare** | **Nonessential** | 0.69 | 0.53 | 0.92 | 0.0107 | * | 0.74 | 0.54 | 1.01 | 0.0540 | |
| **Other Essential** | | 0.65 | 0.48 | 0.90 | 0.0077 | * | 0.64 | 0.44 | 0.92 | 0.0162 | * |
| **Furloughed/Job Seeker** | **Employed** | 0.92 | 0.61 | 1.41 | 0.6821 | | 0.96 | 0.60 | 1.56 | 0.8841 | |
| **Unemployed** | | 1.00 | 0.74 | 1.36 | 0.9874 | | 0.86 | 0.61 | 1.22 | 0.3991 | |
| **African American/Black** | **White** | 0.74 | 0.52 | 1.08 | 0.1129 | | 0.58 | 0.38 | 0.91 | 0.0165 | * |
| **Hispanic/Latinx** | | 0.80 | 0.54 | 1.21 | 0.2790 | | 1.07 | 0.62 | 1.84 | 0.8080 | |
| **Asian** | | 2.23 | 0.86 | 7.69 | 0.1432 | | 2.45 | 0.78 | 7.73 | 0.1266 | |
| **Multiracial/Other** | | 0.79 | 0.53 | 1.20 | 0.2465 | | 0.72 | 0.43 | 1.21 | 0.2164 | |
| **1 Preexisting** | **0** | 0.85 | 0.63 | 1.14 | 0.2869 | | 0.99 | 0.69 | 1.43 | 0.9620 | |
| **2 Preexisting** | | 0.88 | 0.64 | 1.21 | 0.4216 | | 1.00 | 0.69 | 1.46 | 0.9995 | |
| **3+ Preexisting** | | 0.87 | 0.62 | 1.22 | 0.4198 | | 1.05 | 0.70 | 1.60 | 0.8072 | |
| **Preexisting Not Say** | | 0.41 | 0.17 | 1.11 | 0.0561 | | 0.95 | 0.22 | 4.14 | 0.9412 | |
| **Parent** | **Not a Parent** | 0.78 | 0.60 | 1.01 | 0.0584 | | 0.63 | 0.45 | 0.89 | 0.0086 | * |
| **Income 0-40K** | **100K+** | 0.81 | 0.46 | 1.44 | 0.4642 | | 0.79 | 0.39 | 1.62 | 0.5221 | |
| **Income 40-70K** | | 0.53 | 0.37 | 0.76 | 0.0007 | ** | 0.56 | 0.37 | 0.85 | 0.0066 | * |
| **Income 70-100K** | | 0.64 | 0.44 | 0.91 | 0.0139 | * | 0.63 | 0.42 | 0.96 | 0.0316 | * |
| **PopDensity 0-149** | **1000+** | 0.48 | 0.35 | 0.67 | 1.82E-05 | *** | 0.53 | 0.34 | 0.82 | 0.0049 | * |
| **PopDensity 150-999** | | 0.79 | 0.61 | 1.01 | 0.0581 | | 0.81 | 0.59 | 1.11 | 0.1841 | |
| **West** | **Northeast** | 0.87 | 0.50 | 1.49 | 0.6039 | | 0.95 | 0.50 | 1.79 | 0.8640 | |
| **Midwest** | | 0.75 | 0.46 | 1.22 | 0.2380 | | 0.73 | 0.43 | 1.23 | 0.2372 | |
| **South** | | 0.62 | 0.38 | 1.01 | 0.0563 | | 0.69 | 0.40 | 1.17 | 0.1669 | |
| **Practiced Protective Behavior** | **Did Not Practice** | 1.17 | 0.91 | 1.53 | 0.2209 | | 1.06 | 0.79 | 1.41 | 0.7170 | |
| **State Cumulative Case Rate** | | 0.95 | 0.85 | 1.06 | 0.3754 | | 0.99 | 0.86 | 1.13 | 0.8526 | |
| **State Cumulative Death Rate** | | 0.83 | 0.54 | 1.28 | 0.4071 | | 0.90 | 0.55 | 1.46 | 0.6601 | |

| | | | | | | | | | | | |
|---|---|---|---|---|---|---|---|---|---|---|---|
| **Average Daily Pos Test Rate (Nov.)** | | 0.98 | 0.94 | 1.02 | 0.3269 | | 0.97 | 0.92 | 1.02 | 0.1763 | |
| **County % without HS Degree** | | 1.00 | 0.97 | 1.02 | 0.8621 | | 0.99 | 0.96 | 1.02 | 0.6456 | |
| **Received COVID Test** | **Never Received Test** | 1.00 | 0.73 | 1.39 | 0.9845 | | 1.05 | 0.75 | 1.49 | 0.7635 | |
| **Vaccine Intent - Unlikely** | **Vaccine Intent - Likely** | 0.02 | 0.01 | 0.02 | 2.25E-183 | *** | 0.02 | 0.01 | 0.03 | 2.07E-114 | *** |
| **Vaccine Intent - Undecided** | **Vaccine Intent - Likely** | 0.08 | 0.06 | 0.11 | 1.22E-67 | *** | 0.08 | 0.06 | 0.12 | 1.06E-39 | *** |

*: $p < 0.05$
**: $p < 0.001$
***: $p < 0.0001$

**Supplementary Table 13: Hesitant User Analysis** - Weighted vaccine uptake model for only users that responded as "hesitant" to the vaccine intent question.

| | | Vaccine Uptake Analysis for Hesitant Users (n = 2,520) | | | | |
|---|---|---|---|---|---|---|
| **Variable** | **Reference Group** | **OR** | **Lower 95% CI** | **Upper 95% CI** | **P-Value** | **Sig** |
| **Female** | **Male** | 0.68 | 0.44 | 1.06 | 0.0894 | |
| **Age [18,30)** | **65+** | 0.14 | 0.07 | 0.25 | 5.48E-10 | *** |
| **Age [30,45)** | | 0.27 | 0.16 | 0.45 | 4.79E-07 | *** |
| **Age [45,55)** | | 0.40 | 0.24 | 0.66 | 0.0003 | ** |
| **Age [55,65)** | | 0.60 | 0.37 | 0.99 | 0.0460 | * |
| **Essential Healthcare** | **Nonessential** | 0.66 | 0.47 | 0.94 | 0.0222 | * |
| **Other Essential** | | 0.65 | 0.44 | 0.97 | 0.0336 | * |
| **Furloughed/Job Seeker** | **Employed** | 1.18 | 0.69 | 2.01 | 0.5501 | |
| **Unemployed** | | 1.11 | 0.74 | 1.66 | 0.6231 | |
| **African American/Black** | **White** | 0.76 | 0.50 | 1.15 | 0.1908 | |
| **Hispanic/Latinx** | | 0.98 | 0.57 | 1.68 | 0.9379 | |
| **Asian** | | 3.18 | 0.70 | 14.42 | 0.1337 | |
| **Multiracial/Other** | | 0.80 | 0.46 | 1.38 | 0.4241 | |
| **1 Preexisting** | **0** | 0.97 | 0.67 | 1.39 | 0.8504 | |
| **2 Preexisting** | | 0.87 | 0.59 | 1.29 | 0.4989 | |
| **3+ Preexisting** | | 0.94 | 0.61 | 1.44 | 0.7672 | |
| **Preexisting Not Say** | | 2.34 | 0.61 | 8.98 | 0.2158 | |
| **Parent** | **Not a Parent** | 0.79 | 0.57 | 1.10 | 0.1565 | |
| **Income 0-40K** | **100K+** | 1.15 | 0.55 | 2.38 | 0.7143 | |
| **Income 40-70K** | | 0.60 | 0.39 | 0.90 | 0.0149 | * |
| **Income 70-100K** | | 0.80 | 0.52 | 1.23 | 0.3099 | |
| **PopDensity 0-149** | **1000+** | 0.48 | 0.31 | 0.75 | 0.0012 | * |
| **PopDensity 150-999** | | 0.79 | 0.58 | 1.08 | 0.1428 | |
| **West** | **Northeast** | 0.76 | 0.39 | 1.51 | 0.4392 | |
| **Midwest** | | 0.75 | 0.41 | 1.37 | 0.3510 | |
| **South** | | 0.66 | 0.36 | 1.20 | 0.1764 | |
| **Practiced Protective Behavior** | **Did Not Practice** | 0.91 | 0.67 | 1.25 | 0.5592 | |
| **State Cumulative Case Rate** | | 0.93 | 0.80 | 1.08 | 0.3527 | |
| **State Cumulative Death Rate** | | 0.79 | 0.46 | 1.36 | 0.3954 | |
| **Average Daily Pos Test Rate (Nov.)** | | 0.99 | 0.93 | 1.04 | 0.6154 | |
| **County % without HS Degree** | | 0.99 | 0.96 | 1.03 | 0.6810 | |
| **Received COVID Test** | **Never Received Test** | 0.90 | 0.60 | 1.35 | 0.6181 | |
| **Vaccine Intent - Undecided** | **Vaccine Intent - Unlikely** | 4.57 | 3.47 | 6.03 | 2.26E-26 | *** |

*: $p < 0.05$
**: $p < 0.001$
***: $p < 0.0001$
